# Reconstructing the SARS-CoV-2 epidemic in eastern Uganda through longitudinal serosurveillance in a malaria cohort

**DOI:** 10.1101/2022.09.20.22280170

**Authors:** Jessica Briggs, Saki Takahashi, Patience Nayebare, Gloria Cuu, John Rek, Maato Zedi, Timothy Kizza, Emmanuel Arinaitwe, Joaniter I. Nankabirwa, Moses Kamya, Prasanna Jagannathan, Karen Jacobson, Philip J. Rosenthal, Grant Dorsey, Bryan Greenhouse, Isaac Ssewanyana, Isabel Rodríguez-Barraquer

## Abstract

**Importance:** Estimating the true burden of SARS-CoV-2 infection has been difficult in sub-Saharan Africa due to asymptomatic infections and inadequate testing capacity. Antibody responses from serologic surveys can provide an estimate of SARS-CoV-2 exposure at the population level.

**Objective:** To estimate SARS-CoV-2 seroprevalence, attack rates, and re-infection in eastern Uganda using serologic surveillance from 2020 to early 2022.

**Design:** Plasma samples from participants in the Program for Resistance, Immunology, Surveillance, and Modeling of Malaria in Uganda (PRISM) Border Cohort were obtained at four sampling intervals: October-November 2020; March-April 2021; August-September 2021; and February-March 2022. <u>Setting</u>: Tororo and Busia districts, Uganda.

**Participants:** 1,483 samples from 441 participants living in 76 households were tested. Each participant contributed up to 4 time points for SARS-CoV-2 serology, with almost half of all participants contributing at all 4 time points, and almost 90% contributing at 3 or 4 time points. Information on SARS-CoV-2 vaccination status was collected from participants, with the earliest reported vaccinations in the cohort occurring in May 2021.

**Main Outcome(s) and Measure(s):** The main outcomes of this study were antibody responses to the SARS-CoV-2 spike protein as measured with a bead-based serologic assay. Individual-level outcomes were aggregated to population-level SARS-CoV-2 seroprevalence, attack rates, and boosting rates. Estimates were weighted by the local age distribution based on census data.

**Results:** By the end of the Delta wave and before widespread vaccination, nearly 70% of the study population had experienced SARS-CoV-2 infection. During the subsequent Omicron wave, 85% of unvaccinated, previously seronegative individuals were infected for the first time, and ∼50% or more of unvaccinated, already seropositive individuals were likely re-infected, leading to an overall 96% seropositivity in this population. Our results suggest a lower probability of re-infection in individuals with higher pre-existing antibody levels. We found evidence of household clustering of SARS-CoV-2 seroconversion. We found no significant associations between SARS-CoV-2 seroconversion and gender, household size, or recent *Plasmodium falciparum* malaria exposure.

**Conclusions and Relevance:** Findings from this study are consistent with very high infection rates and re-infection rates for SARS-CoV-2 in a rural population from eastern Uganda throughout the pandemic.

## INTRODUCTION

Understanding population-level exposure and immunity to SARS-CoV-2 is necessary to measure transmission, determine the susceptible population, and inform public health responses. However, estimating the proportion of the population that has been infected, particularly in resource-limited settings, is complicated by asymptomatic or subclinical infections, inadequate testing capacity, and challenges in collecting routine surveillance data. Seroprevalence surveys can overcome these issues by identifying antibody responses that reflect prior SARS-CoV-2 exposure^1^. A recent review of seroprevalence studies conducted in sub-Saharan Africa when vaccination coverage was low (<5% in September 2021) reported a rise in SARS-CoV-2 seroprevalence from 3% in Q2 2020 to 65% in Q3 2021^2^.

There are little reported SARS-CoV-2 seroprevalence data specific to Uganda to elucidate the true burden of infection and under-ascertainment of cases. Only one population-level serosurvey in Uganda has been published thus far, which identified an overall seroprevalence of 21% in March 2021^3^. To our knowledge, there have also been no published serosurveys in sub-Saharan Africa that estimate population seroprevalence after the Omicron wave or the attack rate of the Omicron variant. Here, we leveraged samples collected as part of a longitudinal malaria cohort study in eastern Uganda to reconstruct the epidemic trajectory of SARS-CoV-2 over the first two years of the pandemic and estimate the attack rate of the main epidemic waves, along with risk factors for seroconversion.

## METHODS

### Study design

This study was conducted within the Program for Resistance, Immunology, Surveillance, and Modeling of Malaria in Uganda (PRISM) Border Cohort study in the Tororo and Busia districts of Uganda. The design and population of this ongoing cohort have recently been described elsewhere^4^. At cohort study enrollment, a survey was conducted to collect information on household characteristics, including household wealth tertile^5^, sanitation category, and housing type (traditional versus modern).

Participants from the 80 households enrolled in the cohort were encouraged to come to a dedicated study clinic open 7 days per week for all medical care which they received free of charge. Routine study visits were conducted every 4 weeks and included a standardized evaluation and blood collection. Information on SARS-CoV-2 vaccination status and dates was collected from participants.

For this SARS-CoV-2 study, we leveraged the biospecimen repository of plasma samples collected at monthly routine study visits. We selected four sampling intervals between October 2020 and March 2022, informed by COVID-19 case counts in Uganda (**Figure 1A** and **Supplementary Figure 1**). For sampling, we included all available samples from study participants who had a routine visit during the Round 3 sampling interval.

**Figure 1:**
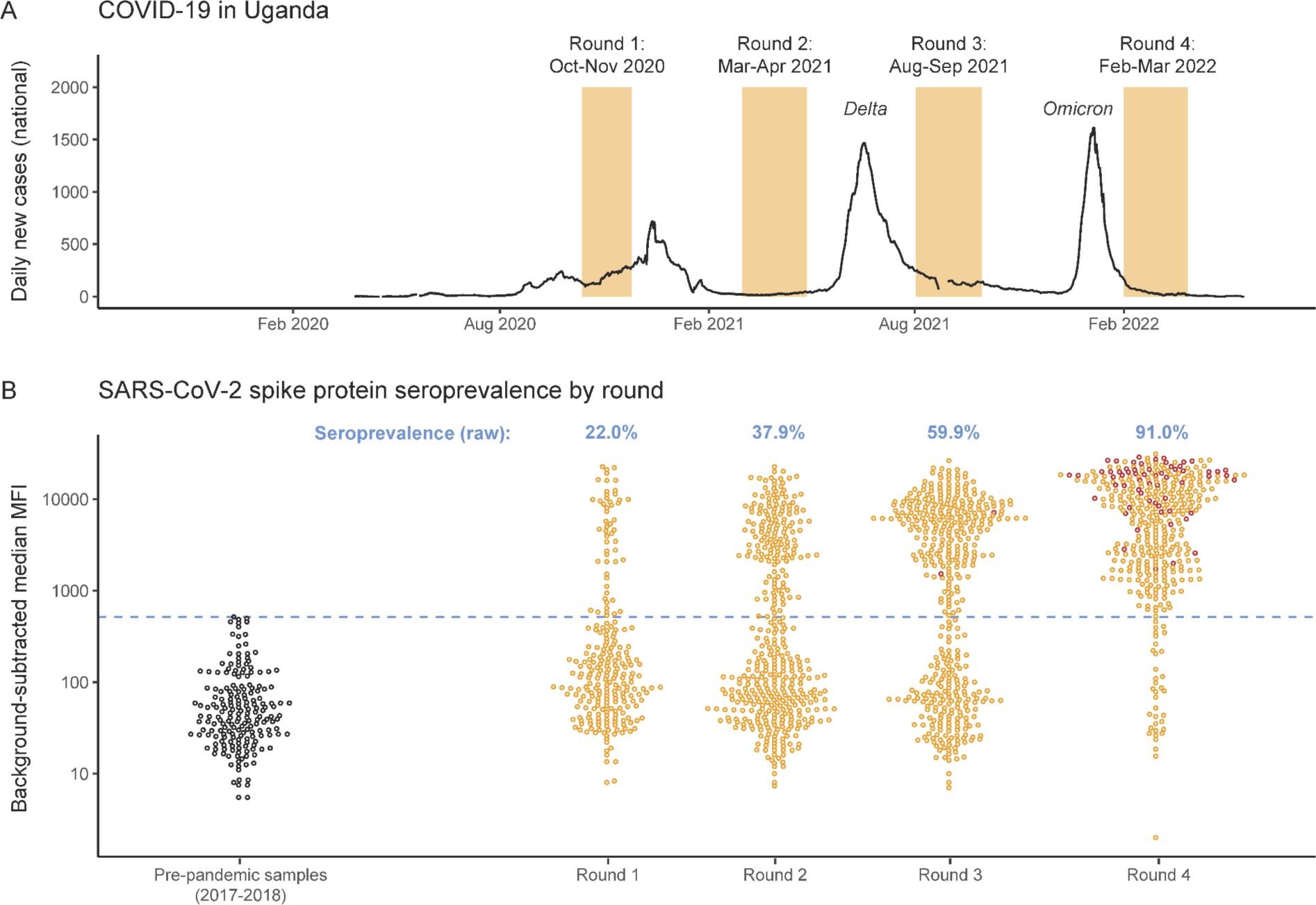
The SARS-CoV-2 epidemic in Uganda and in the PRISM Border Cohort study. **(A)** Reported daily new COVID-19 cases in Uganda (seven-day rolling average), obtained from the Our World in Data database: https://ourworldindata.org/coronavirus/country/uganda. One week in late August 2021 where >21,000 cases were reported in one day is omitted. The four rounds of the PRISM Border Cohort SARS-CoV-2 serosurveys are shown in orange. **(B)** SARS-CoV-2 seroprevalence by round. The y-axis shows the background-subtracted median MFI of the spike protein antibody response. The left-most beeswarm plot (black circles) shows responses from 192 pre-pandemic control samples from the PRISM-2 study (also in Tororo District, Uganda), collected in 2017-2018. The subsequent beeswarm plots show the distribution of antibody responses by serosurvey round. Visits from participants who had received SARS-CoV-2 vaccination by 3 weeks before the serosurvey sample (i.e., to allow for seroconversion after vaccination) are shown as dark red circles. The cutoff for seropositivity is shown in the blue dashed line (background-subtracted median MFI = 516). The raw seroprevalence by round, not adjusted for test performance characteristics or covariates, is shown in blue text above each beeswarm plot.

The study protocol was approved by the Makerere University School of Medicine Research and Ethics Committee, the Uganda National Council of Science and Technology, the University of California, San Francisco, Human Research Protection Program, the London School of Hygiene & Tropical Medicine Ethics Committee, and the Stanford University Research Compliance Office. Written informed consent was obtained for all participants prior to study enrollment.

### Laboratory methods

A multiplex bead assay and protocol previously described for SARS-CoV-2 serology studies^6^,^7^ was modified to assess total IgG responses to the spike protein, the receptor binding domain of the spike protein (RBD), and the nucleocapsid protein. Detailed methods are in the **Supplementary Methods**. Briefly, plasma samples were assayed at a 1:400 dilution to determine antibody seropositivity. The results were expressed as mean fluorescent intensity (MFI). A standard curve using a pool of positive serum was included on each plate to normalize for plate-to-plate variations (**Supplementary Figure 2**) and to infer relative antibody concentrations using a 4-parameter logistic model^8^. Samples run in duplicate demonstrated high replicability (**Supplementary Figure 3**). A subset of samples was tested at a 1:4,000 dilution to assess antibody boosting. A greater dilution for the boosting assay was used, since many seropositive samples were near or outside the assay’s linear dynamic range at 1:400.

For negative controls, we used 192 pre-pandemic plasma samples from the PRISM-2 cohort study also in Tororo District, Uganda^9^, collected in 2017 and 2018 (**Supplementary Figure 4**). These samples were selected to have the same age distribution as the study population and represented an appropriate background for serologic signals (e.g., cross-reactivity from other circulating coronaviruses^10^). For positive controls, we used 156 plasma samples from 99 volunteers enrolled in the UCSF Long-term Impact of Infection with Novel Coronavirus (LIINC) study^6^. Cutoff for seropositivity was established as the highest value among the negative controls, and therefore specificity was 100% (192/192) by design. Assay sensitivity was calculated as the proportion of the positive controls above the cutoff, and was 92.9% (145/156) (**Supplementary Figure 5**). As an additional check on assay performance characteristics, we calculated the sensitivity in samples from 11 participants from the PRISM Border Cohort study who tested positive for symptomatic SARS-CoV-2 infection between February and July 2021 (Mwakibete et al, submitted). 10 of 11 (90.9%) seroconverted, and responses remained robust over time (**Supplementary Figure 6**).

### Statistical analyses

We focused on spike antibody responses and used background-subtracted MFIs as the primary outcome. A sensitivity analysis was performed using relative concentrations as the outcome and demonstrated highly concordant results (**Supplementary Figure 7**). While the assay also measured RBD and nucleocapsid responses, the RBD and spike responses were highly correlated, and the nucleocapsid responses had high background in this assay as previously noted^6^, and thus were not well suited to evaluate primary seroconversion to natural infection (**Supplementary Figure 8**).

Raw SARS-CoV-2 spike seropositivity was determined as the proportion of samples that tested positive at the 1:400 dilution. We then estimated seroprevalence, adjusted for sensitivity and specificity, using a Bayesian measurement model^11^. The attack rate was determined as the proportion of individuals seronegative in one serosurvey round who were seropositive at the subsequent round. We computed 95% credible intervals (CrI) to quantify uncertainty in posterior estimates. We produced estimates weighted by the local age distribution based on 2014 census data from the Uganda Bureau of Statistics (https://www.ubos.org/).

To assess SARS-CoV-2 boosting, we analyzed responses at the 1:4,000 dilution. We limited assessment of boosting to between Rounds 3 and 4 in individuals who were already seropositive at Round 3. In the primary analysis we defined boosting as a ≥ 4 fold increase in response, a commonly used threshold across pathogens for ascertaining recent infection^12^-^14^. In sensitivity analyses, we assessed alternative definitions, i.e., ≥ 2 and ≥ 8 fold increases and incorporating nucleocapsid responses. As participants were receiving vaccinations during this time interval, nucleocapsid responses were included in evaluation of boosting (but not primary seroconversion) as they can potentially differentiate antibody responses resulting from infection versus vaccination with spike-based vaccines. The latter do not generate nucleocapsid responses and include the majority of SARS-CoV-2 vaccines received by participants in this cohort.

To test for individual and household characteristics (and visit-level patient-reported symptoms and clinician-assigned diagnoses) associated with SARS-CoV-2 seroconversion and boosting, univariate binomial regression was performed. For associations with seroconversion, patients could contribute multiple observations until seroconversion; therefore, generalized estimating equations were used to adjust for repeated measures^15^. For associations with boosting, only children were included because SARS-CoV-2 vaccination had begun in those 18 years and older by the time of sample collection. Malaria incidence between each sampling round was calculated by dividing the number of episodes of clinical malaria by the number of days of observation for each individual. Asymptomatic parasitemia was defined as the presence of *P. falciparum* parasites via microscopy or *var*ATS qPCR^16^ and the absence of fever (measured or by history).

To test for clustering of seroconversions within households, we adapted an approach of estimating pairwise odds ratios to detect correlation structure in binary data^17–19^ (see **Supplementary Methods**). All analyses were conducted using the R statistical software (https://www.r-project.org/) and the Stan programming language (https://mc-stan.org/).

## RESULTS

### Clinical and demographic characteristics of the study participants

A total of 1,483 samples from 441 participants living in 76 households were tested (**Table 1**). Participants contributed up to 4 time points (Figure 1A), with almost half of participants contributing at all 4 time points, and almost 90% contributing at 3 or 4 time points. Household sizes ranged from 3 to 8 residents. Results had approximately equal representation from those under 5 years of age, between 5 to 15 years of age, and 16 years of age or older at the time of their first sample collection. The earliest reported SARS-CoV-2 vaccinations in the cohort occurred in May 2021: 3 adults were vaccinated between Round 2 (March-April 2021) and Round 3 (August-September 2021), and 105 of 137 adults (77%) included in Round 4 (February-March 2022) received SARS-CoV-2 vaccination between Rounds 3 and 4 (**Supplementary Figure 9**). 1 participant under 15 years received SARS-CoV-2 vaccination at 2 weeks before their Round 4 sample. While SARS-CoV-2 diagnostic testing was not systematically performed, 7 participants reported having tested positive by RDT. There were no hospitalizations or deaths due to COVID-19 in the cohort.

**Table 1:** Demographic characteristics of the study participants.

| **Participants (N = 441)** |  |
| --- | --- |
| Age at first time point included | < 5 years: 156 (35.4%)<br>5-15 years: 130 (29.5%)<br>16 years or older: 155 (35.1%) |
| Sex | Female: 245 (55.6%)<br>Male: 196 (44.4%) |
| District | Tororo: 350 (79.4%)<br>Busia: 91 (20.6%) |
| **Households (N = 76)** |  |
| Number of participants from a household | 3: 2 (2.6%)<br>4: 12 (15.8%)<br>5: 16 (21.1%)<br>6: 17 (22.4%)<br>7: 27 (35.5%)<br>8: 2 (2.6%) |
| **Serosurvey rounds (1,483 samples total)** |  |
| Round 1 (Oct 12 to Nov 25, 2020)<br>Round 2 (Mar 2 to Apr 28, 2021)<br>Round 3 (Aug 2 to Sep 29, 2021)<br>Round 4 (Feb 1 to Mar 29, 2022) | 245 samples<br>414 samples<br>434 samples<br>390 samples |
| Number of time points included per participant | 1: 4 (0.9%)<br>2: 46 (10.4%)<br>3: 177 (40.1%)<br>4: 214 (48.5%) |

### SARS-CoV-2 seroprevalence, attack rates, and boosting

The raw seropositivity to spike was 22.0% during Round 1 (October-November 2020, half a year since the first reported case in Uganda) and increased over time, reaching 91.0% seropositivity during Round 4 (post-first Omicron wave) (**Figure 1B**; **Supplementary Table 1**). Weighting by the age distribution of the population and accounting for test performance characteristics, seroprevalence increased from 25.6% (95% CrI: 19.6%, 32.0%) at Round 1, to 43.7% (95% CrI: 38.6%, 48.8%) at Round 2, to 67.7% (95% CrI: 62.5%, 72.6%) at Round 3, to 96.0% (95% CrI: 93.4%, 97.9%) at Round 4 (**Figure 2A**). The weighted seroprevalence among unvaccinated individuals was 68.2% (95% CrI: 62.7%, 73.4%%) in Round 3 and 94.7% (95% CrI: 88.3%, 97.7%) in Round 4.

**Figure 2:**
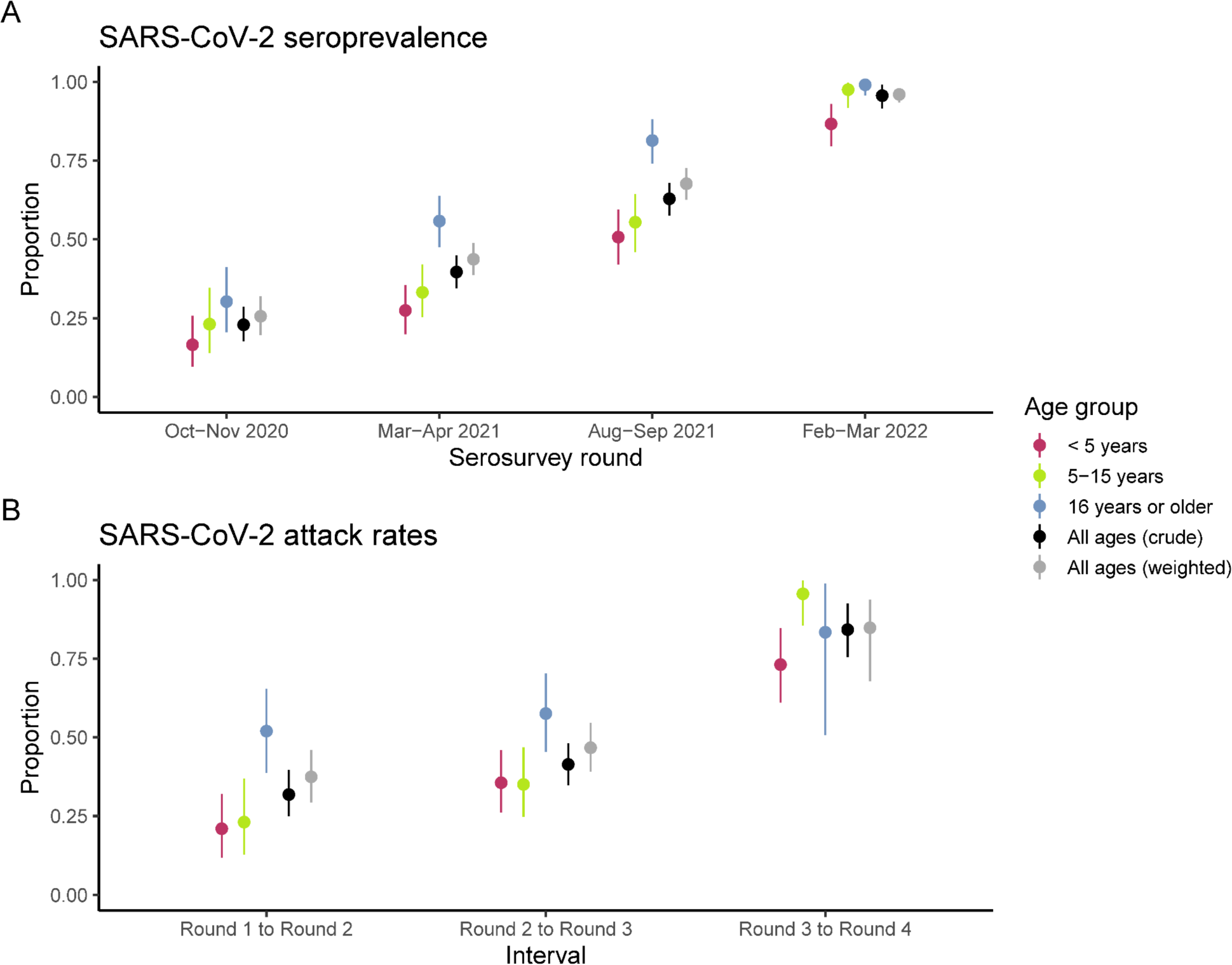
SARS-CoV-2 seroprevalence and attack rates by age group & serosurvey round. **(A)** Posterior median and 95% credible intervals for seroprevalence based on spike protein MFIs, accounting for test performance characteristics. **(B)** Posterior median and 95% credible intervals for attack rate based on spike protein MFIs. Individuals who had received SARS-CoV-2 vaccination by a serosurvey round are removed from the attack rate estimation (e.g., individuals who were vaccinated at Round 4 are removed from the attack rate estimation between Round 3 and Round 4). The colors represent age group-specific estimates. The black values represent the crude estimates in the cohort. The gray values represent estimates weighted by the local age distribution using 2014 census data from the three parishes in Uganda in which study participants reside. All estimates are adjusted for test performance characteristics.

We used longitudinal antibody responses to identify seroconversions (**Supplementary Figure 10**) and to estimate the population-level attack rate of the main epidemic waves (**Figure 2B**; **Supplementary Table 2**). The age-weighted attack rate of the Delta wave (Round 2 to Round 3) was estimated to be 46.7% (95% CrI: 39.1%, 54.6%). The age-weighted attack rate of the Omicron wave (Round 3 to Round 4), omitting individuals who were vaccinated at Round 4, was estimated to be 84.8% (95% CrI: 67.9%, 93.7%).

Among the 232 participants who were seropositive at Round 3 and had a Round 4 sample, we looked for evidence of antibody boosting between these rounds. 125/232 participants (53.9%) boosted based on our primary definition of ≥ 4 fold increase in MFI (**Supplementary Table 3**). This included 70/84 participants vaccinated by Round 4 and 55/148 who had not been vaccinated, consistent with re-infection. Excluding vaccine recipients, and weighting by the age distribution of the population, we estimated that at least 50.8% (95% CrI: 40.6%, 59.7%) of individuals may have been re-infected during the Omicron wave.

Interestingly, we observed more boosting events among individuals who had lower antibody responses at Round 3 than among individuals with higher responses (**Figure 3**). This was consistent across vaccination status, but was particularly evident among unvaccinated individuals. While boosting was observed among 29/41 (71%) of unvaccinated individuals with antibody levels in the lowest tertile, it was only observed in 7/62 (11%) of unvaccinated individuals with antibody levels in the highest tertile. In addition to tertile, boosting risk increased by age (**Supplementary Figure 11**). Alternate estimates of boosting using different thresholds and incorporating nucleocapsid responses are shown in **Supplementary Figure 12** and **Supplementary Figure 13**.

**Figure 3:**
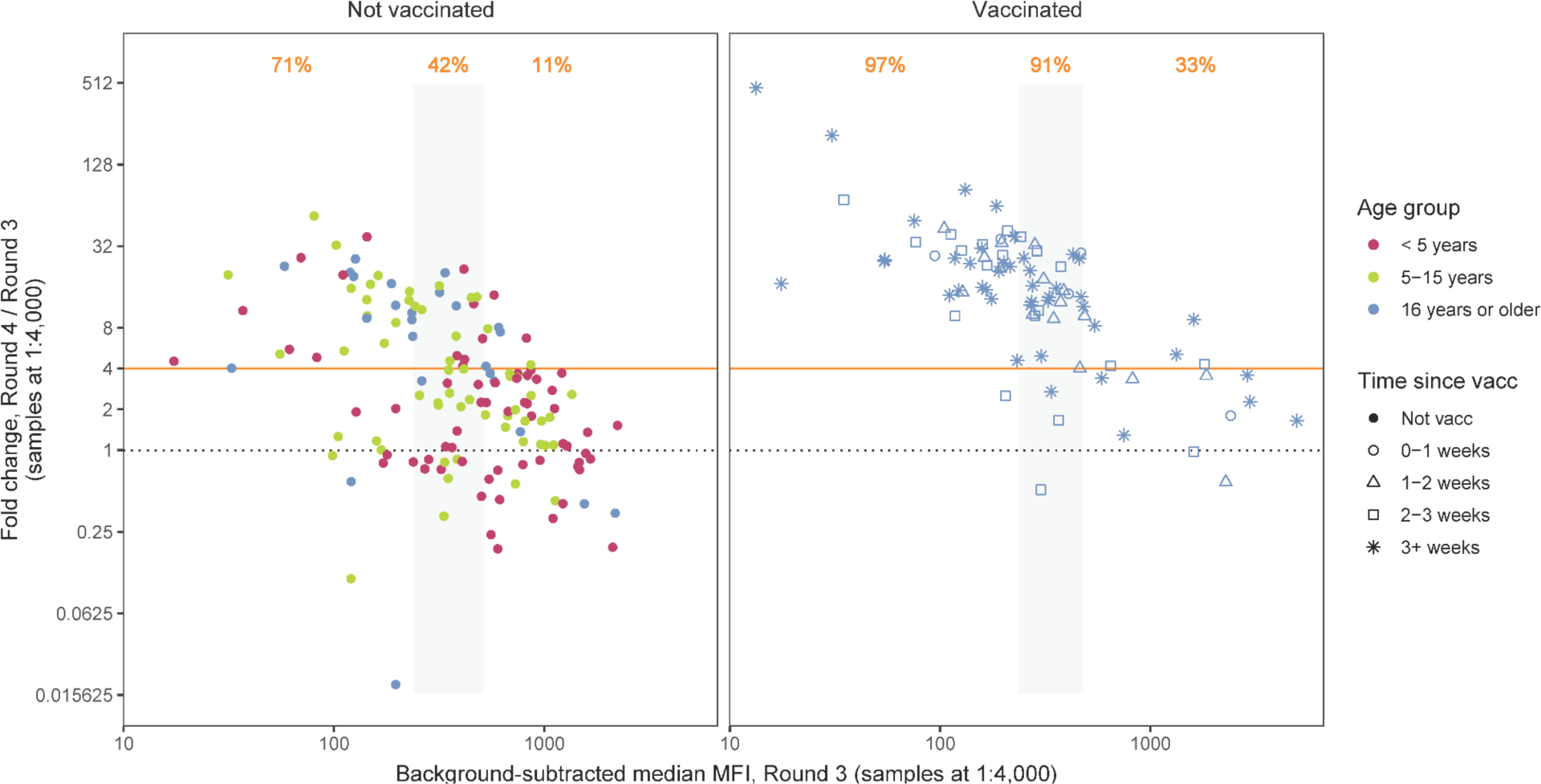
SARS-CoV-2 antibody boosting between Rounds 3 and 4. Round 4 antibody responses among the 232 participants who were seropositive at Round 3 (65 participants < 5 years of age, 58 participants 5-15 years of age, and 109 participants 16 years of age or older) are shown. The Round 3 spike protein antibody response is shown on the x-axis, and the fold change between the Round 4 and Round 3 spike antibody antibody responses is shown on the y-axis. We defined boosting as a ≥ 4 fold increase (orange line). Participants were separated by vaccination status at Round 4 (panels) and by tertiles of Round 3 response (the second tertile is shown in the gray shaded rectangle). The proportion of individuals within each tertile that demonstrated antibody boosting is shown in orange text at the top. The colors of the points represent age groups, and the shapes of the points represent binned time since SARS-CoV-2 vaccination at the Round 4 sample. Note that in the right panel, boosting was observed in 41 of 47 participants vaccinated more than 3 weeks before their Round 4 sample was collected.

### Individual and household level risk factors for SARS-CoV-2 seroconversion and boosting

In univariate analyses, older age was associated with SARS-CoV-2 seroconversion with OR 1.77 (95% confidence interval [CI]: 1.15, 2.74) for adults 16 years and older, and OR 1.31 (95% CI: 0.89, 1.94) for children 5-15 years compared to children less than 5 years (**Table 2**). At each round, seroprevalence was highest among adults, followed by older children, and lowest in younger children (**Figure 2A**). Attack rates were considerably higher in adults during the first two intervals, but not the final interval (**Figure 2B**). We did not find an association between gender and seroconversion.

**Table 2:**
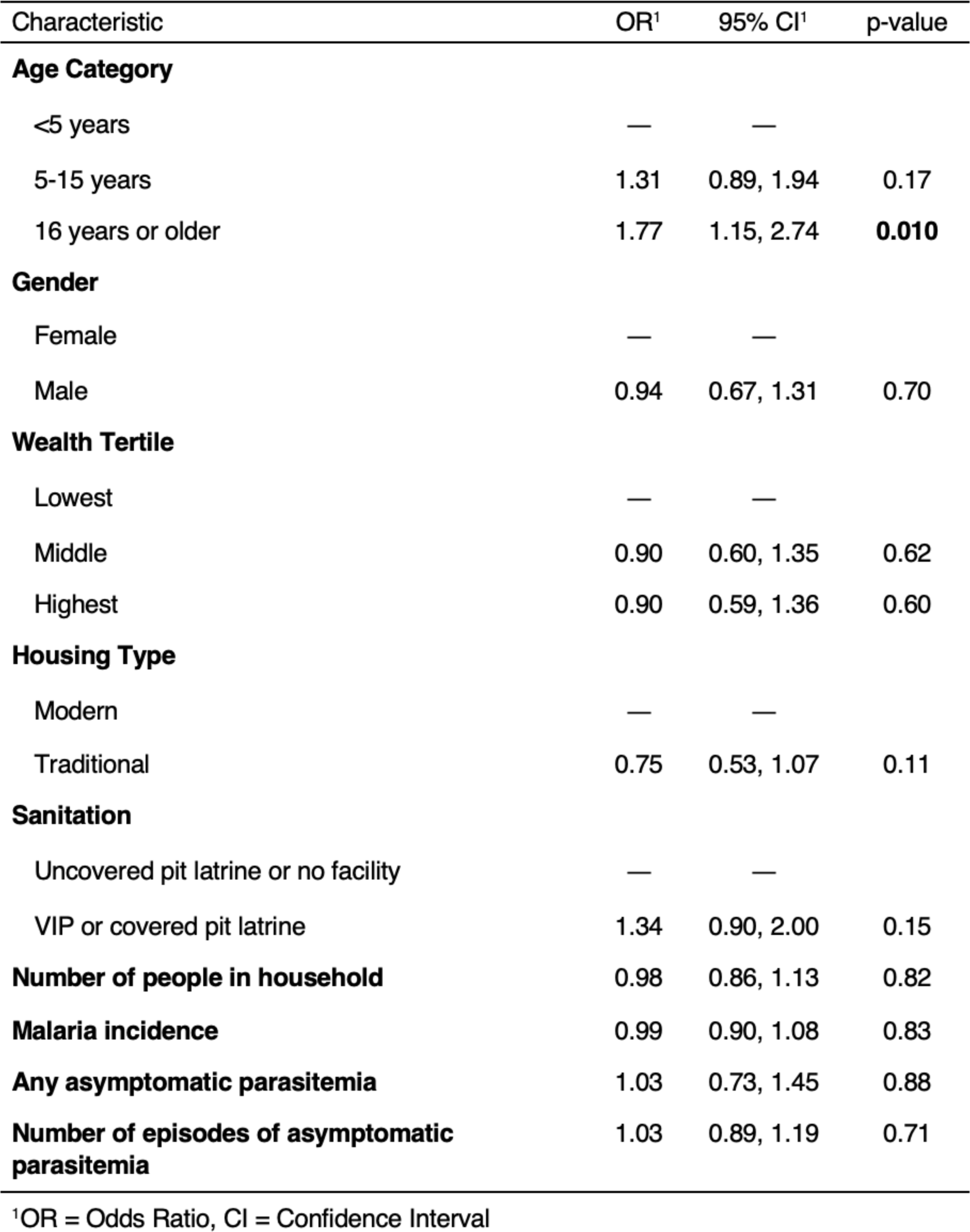
Individual and household level risk factors for SARS-CoV-2 seroconversion.

| Characteristic | OR<sup>1</sup> | 95% CI<sup>1</sup> | p-value |
| --- | --- | --- | --- |
| **Age Category** |  |  |  |
| <5 years | — | — |  |
| 5-15 years | 1.31 | 0.89, 1.94 | 0.17 |
| 16 years or older | 1.77 | 1.15, 2.74 | **0.010** |
| **Gender** |  |  |  |
| Female | — | — |  |
| Male | 0.94 | 0.67, 1.31 | 0.70 |
| **Wealth Tertile** |  |  |  |
| Lowest | — | — |  |
| Middle | 0.90 | 0.60, 1.35 | 0.62 |
| Highest | 0.90 | 0.59, 1.36 | 0.60 |
| **Housing Type** |  |  |  |
| Modern | — | — |  |
| Traditional | 0.75 | 0.53, 1.07 | 0.11 |
| **Sanitation** |  |  |  |
| Uncovered pit latrine or no facility | — | — |  |
| VIP or covered pit latrine | 1.34 | 0.90, 2.00 | 0.15 |
| **Number of people in household** | 0.98 | 0.86, 1.13 | 0.82 |
| **Malaria incidence** | 0.99 | 0.90, 1.08 | 0.83 |
| **Any asymptomatic parasitemia** | 1.03 | 0.73, 1.45 | 0.88 |
| **Number of episodes of asymptomatic parasitemia** | 1.03 | 0.89, 1.19 | 0.71 |

<sup>1</sup>OR = Odds Ratio, CI = Confidence Interval

No household characteristics evaluated were associated with seroconversion (**Table 2**). However, we found evidence of clustering of SARS-CoV-2 seroconversions within households (**Supplementary Figure 14**). Pooled across time intervals and age groups, the odds of seroconversion were 4.33 (95% CrI: 3.27, 5.79) times greater in individuals from households where another individual seroconverted during the same period, as compared to households where other individuals did not seroconvert. Clustering was strongest between child-child pairs (OR 6.42, 95% CrI: 4.31, 9.65), followed by child-adult pairs (OR 2.92, 95% CrI: 1.85, 4.64), and lowest in adult-adult pairs (OR 2.45, 95% CrI: 0.77, 8.06), implying that two children in a household were more likely to seroconvert in the same interval than two adults. These findings suggest an elevated within-household secondary attack rate, particularly amongst children^20^.

To assess potential association between SARS-CoV-2 infection and recent *P. falciparum* infection, we examined the relationship between recent symptomatic malaria and asymptomatic parasitemia with SARS-CoV-2 seroconversion. Malaria incidence and number of episodes of asymptomatic parasitemia were calculated for each SARS-CoV-2 seronegative individual within each time period between sampling rounds. There was no association between an individual’s incidence of symptomatic malaria or occurrence of asymptomatic parasitemia between sampling round and SARS-CoV-2 seroconversion (**Table 2**).

We explored associations between self-reported symptoms and clinician-assigned diagnoses with SARS-CoV-2 seroconversion during a time interval. Symptoms associated with SARS-CoV-2 seroconversion included cough, headache, and fatigue (**Supplementary Table 4**). The number of sick visits between sampling rounds, any upper respiratory tract infection (URTI) diagnosis, and total number of URTIs were also associated with seroconversion.

Finally, these individual and household characteristics were investigated for association with boosting of the SARS-CoV-2 antibody response between Rounds 3 and 4 in children (**Supplementary Table 5**); findings were similar to those in the seroconversion analysis. However, there was an association between antibody boosting and total number of asymptomatic malaria episodes in the time period between Round 3 and Round 4 sampling (OR 1.37, 95% CI: 1.05, 1.81); this association remained but was less strong after adjusting for age (OR 1.29, 95% CI: 0.97, 1.73). There were no specific symptoms or diagnoses associated with increased odds of antibody boosting.

## DISCUSSION

Findings from this study are consistent with very high attack rates of SARS-CoV-2 infection in this rural population from eastern Uganda throughout the pandemic. By the end of the SARS-CV-2 Delta wave and before the widespread availability of vaccination, nearly 70% of the study population had experienced SARS-CoV-2 infection. Moreover, during the subsequent Omicron wave in early 2022, 85% of unvaccinated, previously seronegative individuals were infected for the first time, and at least 51% of unvaccinated, already seropositive individuals were likely re-infected. Based on the SARS-CoV-2 cases reported in Uganda during the time period of this study (163,878 through March 29, 2022) and our estimated weighted seroprevalence in unvaccinated individuals (95% at the end of Round 4), we estimate that only about 1 in 270 infections were ascertained by the reporting system.

The availability of longitudinal sampling enabled us to assess seroconversions and re-infections over time. Interestingly, our results based on serologic evidence of boosting suggested a lower probability of re-infection in individuals with higher pre-existing antibody levels; similar patterns have previously been described in a triple-vaccinated population^21^. Our observation could be explained by several potential factors including immunity (i.e., more effective viral neutralization among individuals with higher pre-existing antibody titers), infection histories with different variants which may affect subsequent boosting^22^, antibody homeostasis (i.e., a ceiling for antibody titers), or limited dynamic range of the assay. Of these, we can only rule out the last explanation, as antibody responses in individuals who did not boost were far from the top of the dynamic range of this assay. If immunity is indeed a driver of this observation, it underscores the importance of achieving and maintaining sufficiently high antibody titers in the population, i.e., through vaccination and revaccination at an appropriate frequency.

The fact that we only tested samples obtained at ∼5 month intervals, and therefore do not know the exact timing of SARS-CoV-2 infection, limits our capacity to make precise inferences about associations between SARS-CoV-2 infection and clinical presentation. Similarly, we could not directly test whether *P. falciparum* infection status affects the risk of SARS-CoV-2 infection or disease. However, we were able to examine whether recent *P. falciparum* infection history was associated with SARS-CoV-2 seroconversion. We found no definitive evidence of an association between recent *P. falciparum* infection history and SARS-CoV-2 infection, despite a weak association between asymptomatic parasitemia and boosting during the Omicron wave in individuals under 16 years of age. Understanding potential immunologic interactions between these pathogens remains an important area of inquiry^23^, particularly in this setting where exposures to the malaria parasite are high^4^.

Our results suggest a higher attack rate during the first Omicron wave of SARS-CoV-2 than the Delta wave (85% vs 47%), and the high estimated re-infection rate during Omicron (51%) is consistent with the evidence of increased infectivity and immune evasion associated with this variant^24,25^. Due to the antigenic changes in the spike protein of the Omicron variant, the sensitivity of existing assays measuring spike antibody responses may be reduced against this variant^26^. Gold standard serologic assays are needed to accurately determine re-infections, possibly with variant-specific assays. This will be increasingly important as re-infections become more frequent and new variants that potentially evade pre-existing immunity continue to emerge. We did not have data to adjust for potential variant-specific reduced sensitivity, nor to incorporate seroreversions (though here and in previous work^6^, we found this assay to demonstrate stable responses over the first few months following infection). Either would lead to increased estimates of seroprevalence, attack rates, and boosting.

These results contribute to a growing body of seroprevalence data from low-resource settings that illustrate very high SARS-CoV-2 infection rates despite low case ascertainment. Infection histories have ultimately generated complex landscapes of SARS-CoV-2 hybrid immunity in different populations. It will be increasingly important to consider additional complexities that have since become central for interpreting the results of SARS-CoV-2 serosurveys, including histories of vaccination, breakthrough infection, and re-infection.

## Data Availability

All data produced in the present study will be made available at time of publication in a peer-reviewed journal, and are available upon reasonable request to the authors.

## ACKNOWLEDGEMENTS

We thank all study participants who took part in this study. We also thank the clinical, laboratory, and data study team and the Infectious Diseases Research Collaboration for administrative and technical support. We acknowledge John Pak and Wesley Wu at the Chan Zuckerberg Biohub for generating the SARS-CoV-2 proteins used in this analysis. We acknowledge Central Public Health Laboratory and Claire Nankoma for sharing district-level COVID-19 case data.

We acknowledge sources of funding support, including from the Schmidt Science Fellows, in partnership with the Rhodes Trust (ST); Chan Zuckerberg Biohub Investigator program (IRB, BG); NIH/NIAID K23AI166009 (JB); NIH/NIAID U19AI089674 (JB, JR, MZ, TK, EA, MK, PJ, GD, BG, IS, IRB); NIH/NIAID U01AI150741 (ST, PJ, BG, IRB); K43TW010365 (JIN); NIH/NIGMS R35GM138361 (IRB).

## COMPETING INTERESTS

The authors have no competing interests to declare.

## SUPPLEMENTARY INFORMATION

### SUPPLEMENTARY METHODS

#### Covalent antigen coupling to magnetic microsphere beads

Antigens were covalently coupled to MagPlex carboxylated magnetic microspheres (Luminex Corp, Austin, USA). 12.5×10^6^ beads for each bead region were suspended by vortexing at medium speed for 2 minutes. The beads were transferred from the stock bottle into a 1.5ml micro-centrifuge tube with low protein binding surface (Eppendorf, UK) to minimize bead loss and centrifuged at 16,000g for 5 minutes. The bead pellet was pulled on the tube side by placing into a magnetic rack for 2 minutes before pipetting off the supernatant. The beads were washed twice by suspension in 1000µl distilled water, vortexed for 30 seconds and centrifuged at 16,000g for 5 minutes. Supernatant was removed after settling the beads on a magnetic rack for 2 minutes. Beads were suspended in 800µl monobasic Sodium Phosphate (NaH_2PO4_, pH 6.2 activation buffer). To activate the carboxyl surface of the beads, 100µl of 50mg/ml Hydroxysulfosuccinimide (Sulfo-NHS) (Thermo Fisher scientific, UK) was added, vortexed briefly and immediately followed by addition of 100µl of 50mg/ml 1-ethyl-3-(3-dimethylaminopropyl) carbodiimide hydrochloride (EDC) (Thermo Fisher scientific, UK). The beads were incubated for 20 minutes at room temperature in the dark, wrapped in aluminum foil on a plate shaker. Beads were pelleted by centrifuging at 16,000g for 5 minutes, placed tubes in a magnetic holder for 2 minutes, the supernatant carefully removed, and washed 3 times with 1000µl PBS. Previously determined amounts of antigen (38.5µl/ml spike, 42ug/ml RBD, and 29.8ug/ml nucleocapsid proteins) were added to each bead region in PBS to make a final volume of 1000µl. The beads/antigen suspension was incubated for 2 hours on a rotating shaker in the dark, covered in aluminum foil. Coupled beads were pelleted by centrifugation at 16,000g for 5 minutes and washed of excess antigen 3 times with 1000µl of phosphate buffered saline 0.05% tween20 (PBS-TBN). The antigen coupled beads were suspended in 1000µl storage buffer (PBS-TBN-Sodium Azide) and stored at 4°C.

#### Assay to measure total IgG

Total IgG responses to spike, RBD, and nucleocapsid proteins were assayed in plasma at 1:400 dilution using the multiplex bead array using the Luminex Magpix machine (Luminex Corp, Austin, Texas). 10ul of each of the three coupled bead regions was added to 5ml PBS-TBN. 50µl of the pooled bead suspension was added to each well, washed and incubated with 50µl of 1:400 test plasma or control sample for 1.5 hours at room temperature. The plates were washed and 50ul of 1/250 goat anti-human IgG rPE labeled antibody (Jackson Immuno Research Laboratories) was added and incubated for 1.5 hours on a shaking platform. The plates were washed, 1xPBS added and read on a MagPix machine.The blank well MFI was deducted from each well to determine the net MFI. We required a minimum of 50 beads per analyte for a measurement to be included in this analysis. Cutoff for seropositivity was established as the highest value among the negative controls; this was determined to be 516 for background-subtracted spike median MFI, and 0.024 for relative spike antibody concentration.

#### Testing for clustering of seroconversions within households using pairwise odds ratios

The pairwise odds ratio is the odds of seroconversion for an individual in a household if another individual from the same household seroconverted, relative to their odds of seroconversion if the individual did not seroconvert. Pairwise odds ratio values greater than 1 indicate clustering within households. We estimated within-age category and between-age category pairwise odds ratios by time interval, and assumed that the pairwise odds ratios were constant across all households. The probabilities and odds were estimated in a multinomial Bayesian framework.

## SUPPLEMENTARY TABLES

**Supplementary Table 1:**
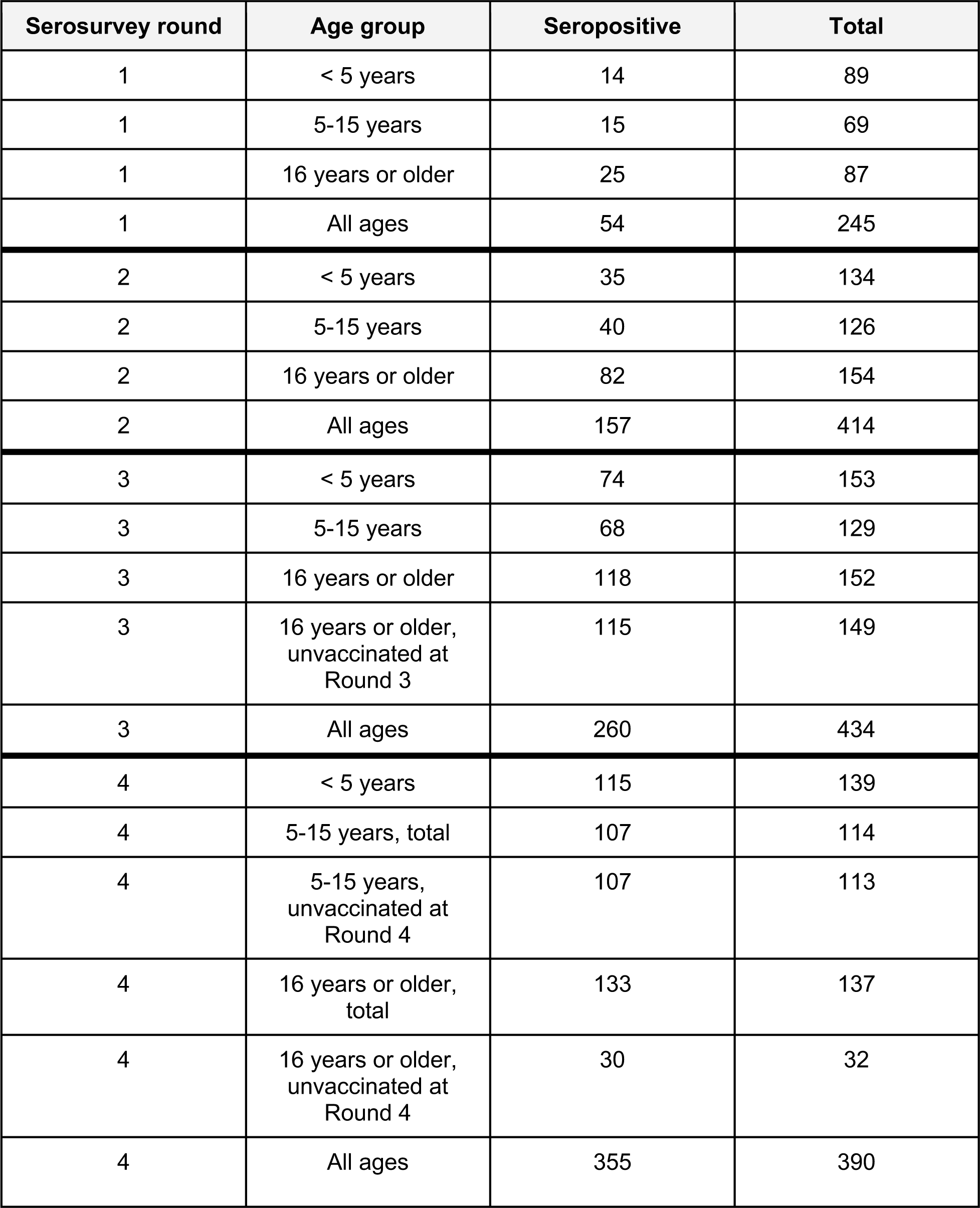
Number of samples seropositive for SARS-CoV-2 by spike protein MFI and total number of samples tested, by serosurvey round and age group.

**Supplementary Table 2:**
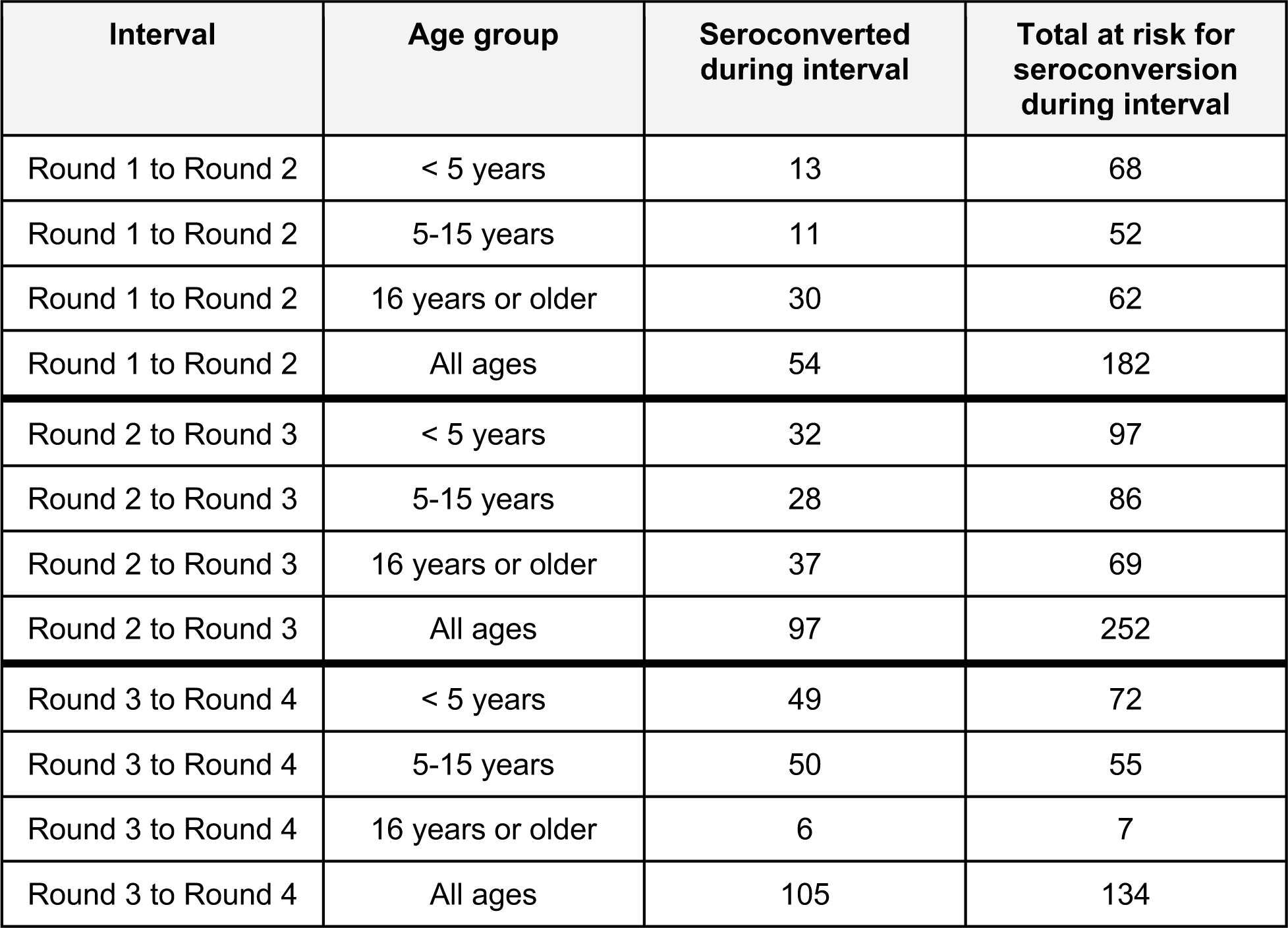
Number of participants with SARS-CoV-2 seroconversion by spike MFI and total number of participants at risk for seroconversion, by interval and age group. Individuals are removed from the risk set if they had received SARS-CoV-2 vaccination before or during the interval.

**Supplementary Table 3:**
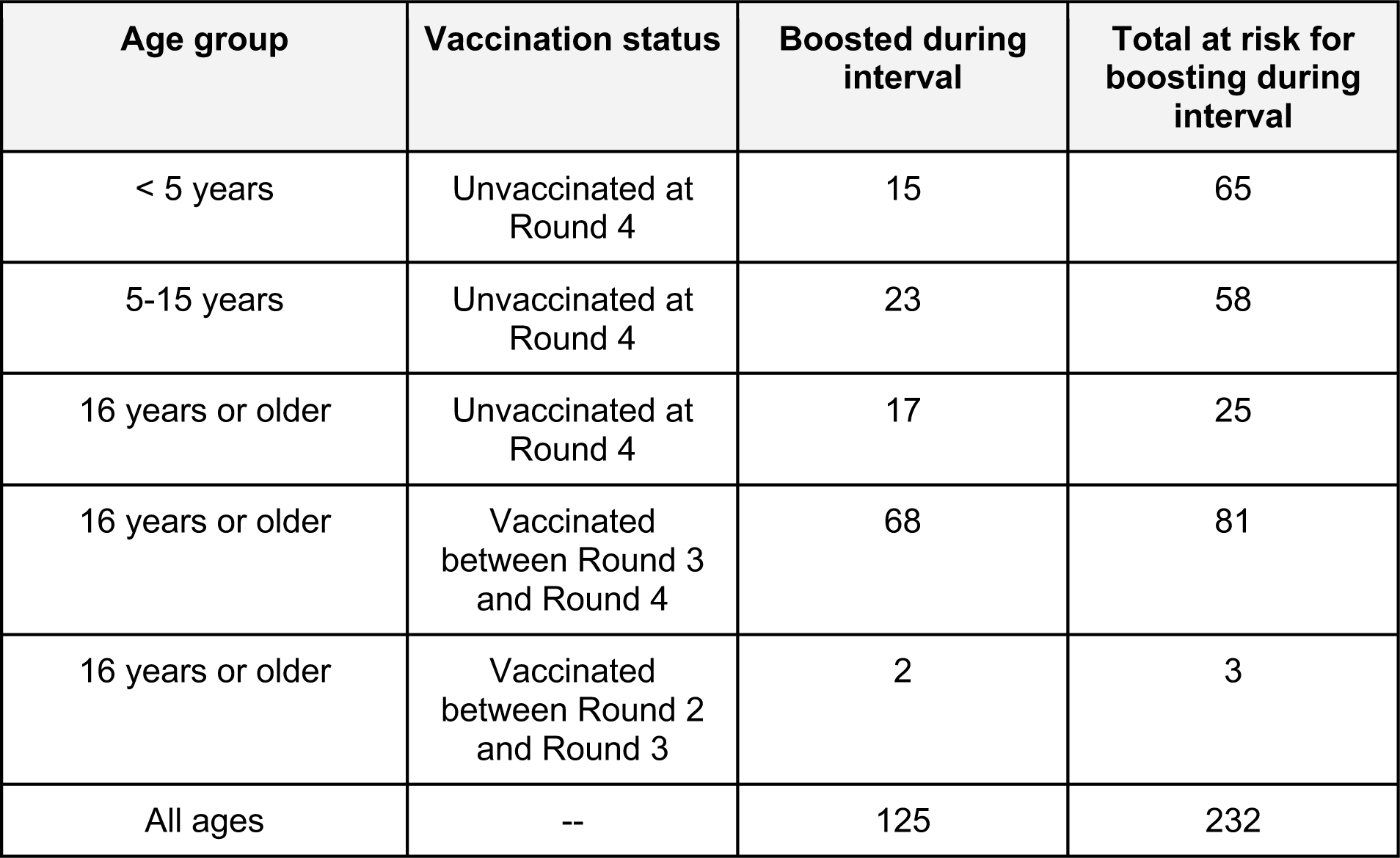
Number of participants with SARS-CoV-2 boosting by spike MFI and total number of participants at risk for boosting, by interval and age group. Only participants who were seropositive at Round 3 were considered at risk for boosting. Boosting was defined as a ≥ 4 fold increase in MFI between Round 3 and Round 4.

**Supplementary Table 4:**
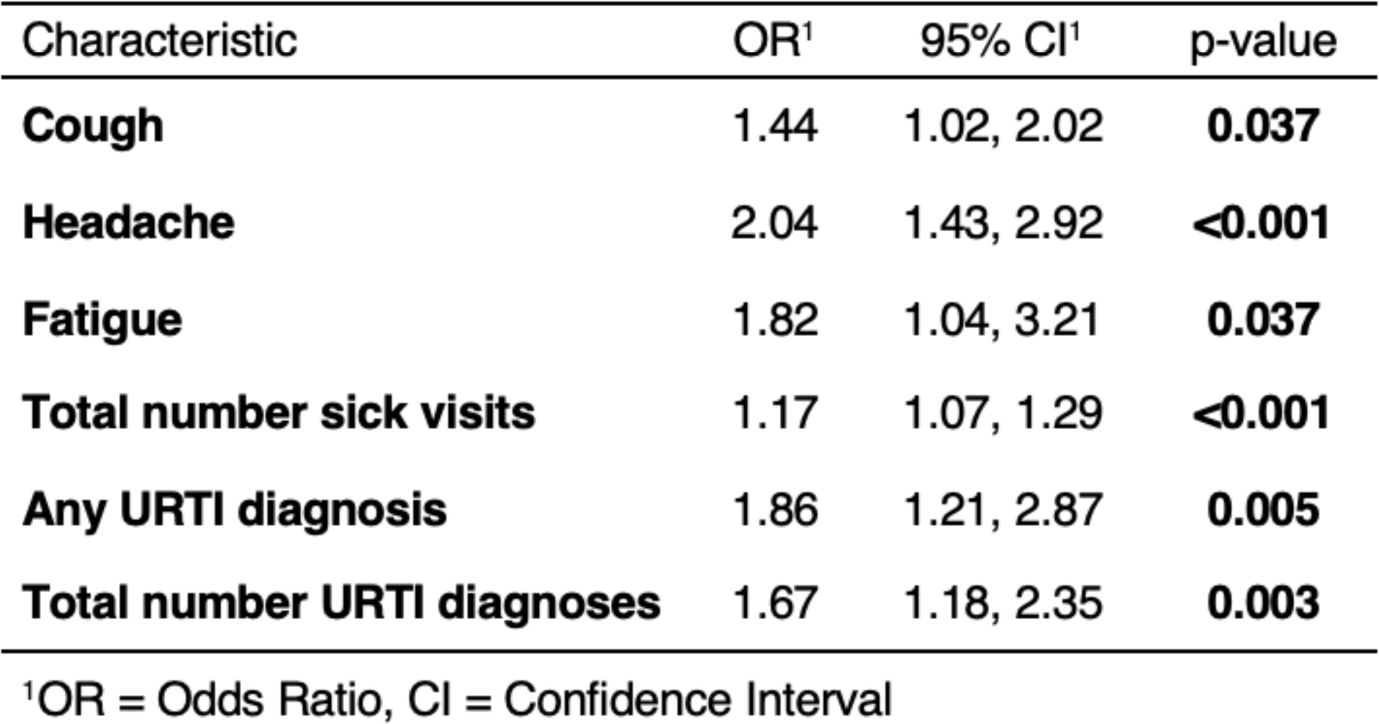
Symptoms and diagnoses associated with SARS-CoV-2 seroconversion. Diarrhea, fever, muscle aches, and other diagnostic categories assessed were not associated with seroconversion.

**Supplementary Table 5:**
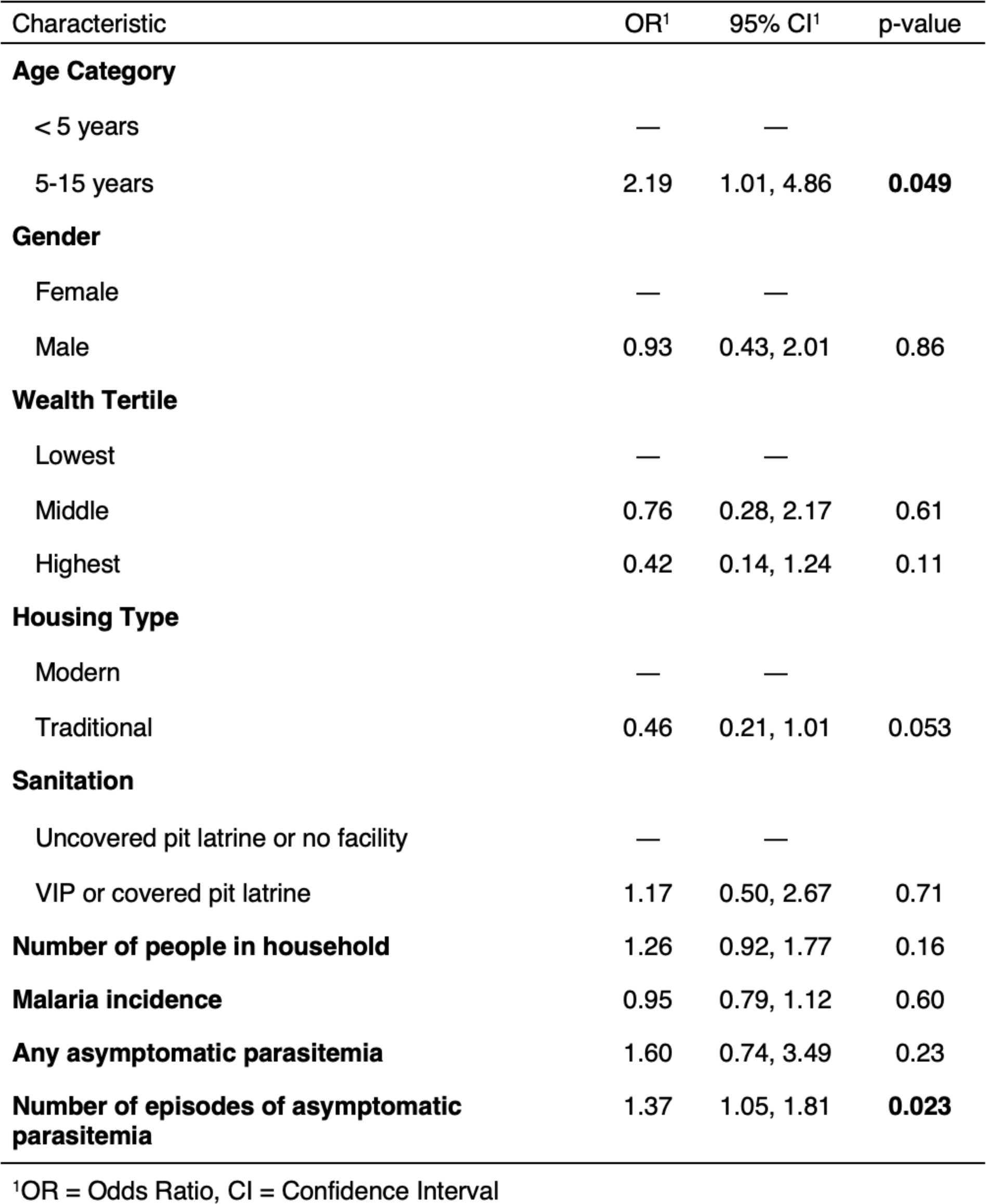
Risk factors for antibody boosting & associations with malaria.

## SUPPLEMENTARY FIGURES

**Supplementary Figure 1:**
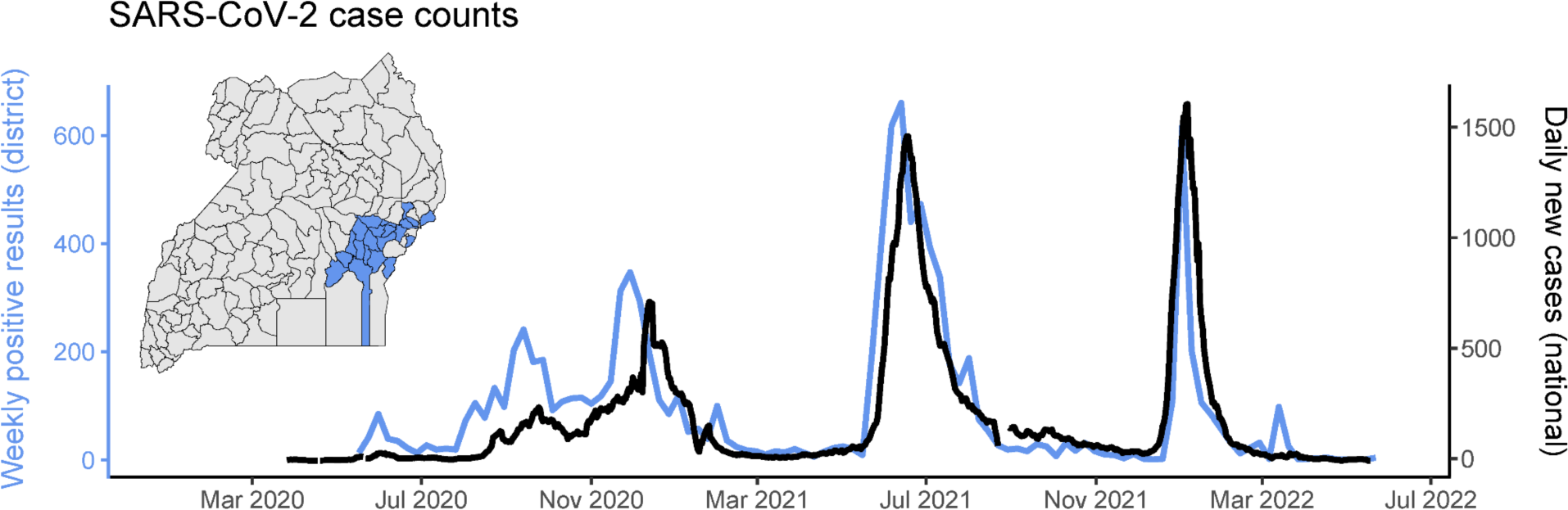
District-level weekly SARS-CoV-2 case counts in eastern Uganda, compared to in the country overall. In blue and on the left y-axis: aggregated weekly SARS-CoV-2 positive results from the 20 districts in eastern Uganda highlighted on the inset map (Budaka, Bugiri, Bugweri, Buikwe, Bukwo, Bulambuli, Busia, Butaleja, Butebo, Iganga, Jinja, Kaliro, Kibuku, Luuka, Mayuge, Mbale, Namisindwa, Namutumba, Pallisa, and Sironko). On the right y-axis: reported daily new COVID-19 cases in all of Uganda, replicated from Figure 1A.

**Supplementary Figure 2:**
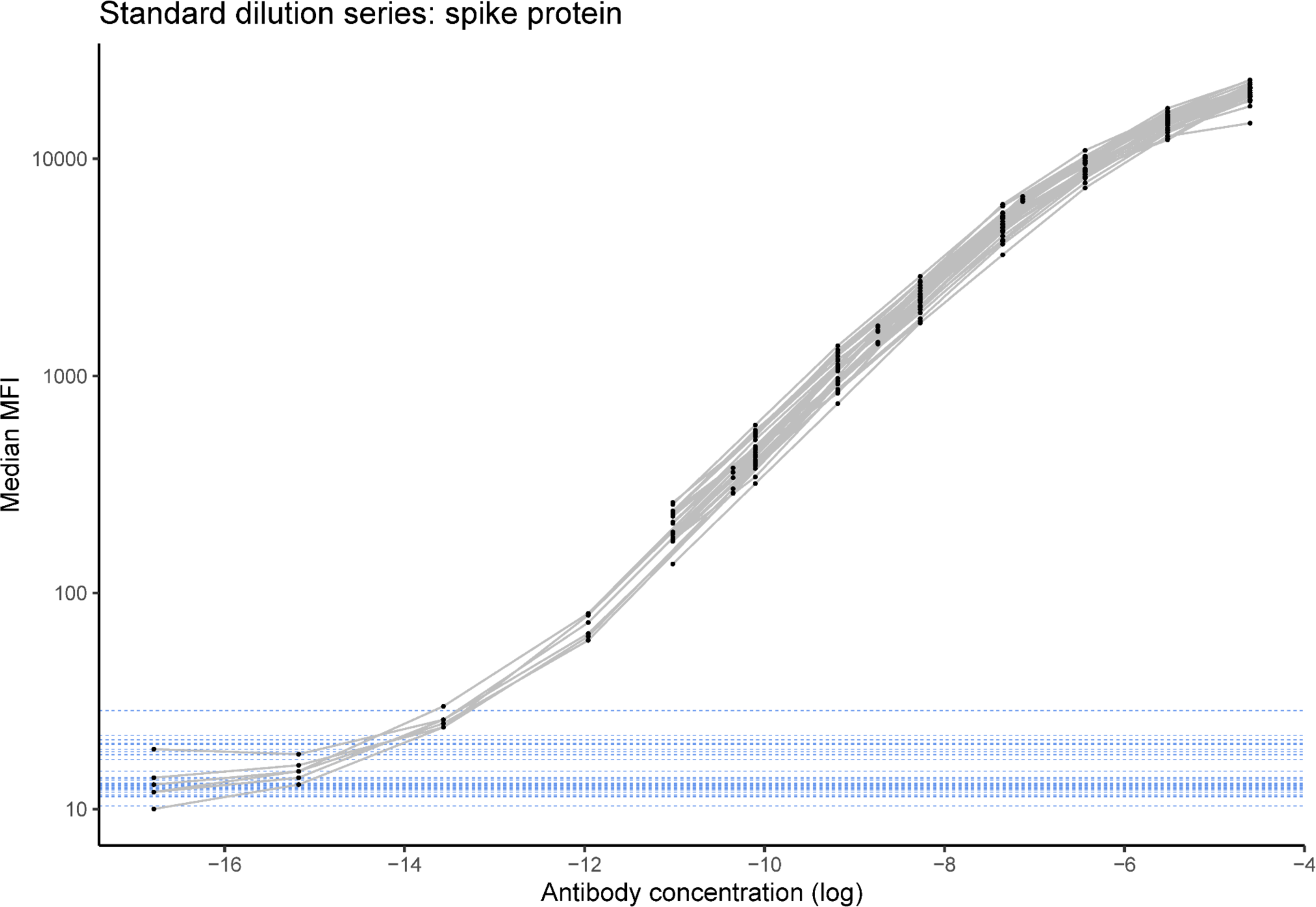
Standard dilution series used for assay normalization. The x-axis represents the log antibody concentration and the y-axis represents the median MFI value of the spike protein response. The black points indicate the serial dilution (inverse of concentration) of the standards. Each line represents a plate, with 32 plates tested in total. The background response (average of blank wells) for each plate is shown in blue.

**Supplementary Figure 3:**
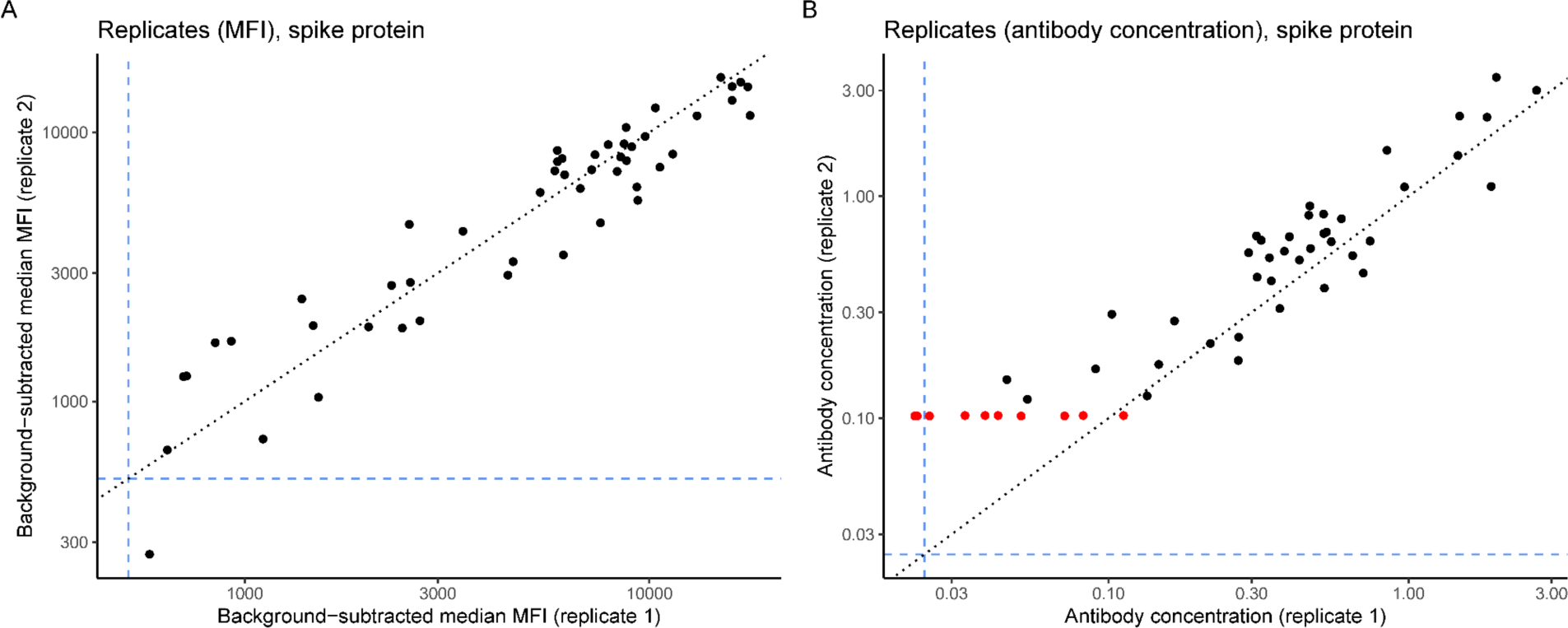
Replicability of the Luminex assay. Scatterplots of spike protein **(A)** MFIs and **(B)** antibody concentrations for 50 samples tested in replicate. The cutoff for seropositivity for each metric, determined based on the highest negative control, is shown in blue. On Panel B, samples that were below the limit of quantitation for antibody concentrations (i.e., below the lowest standard dilution on the plate) are shown in red.

**Supplementary Figure 4:**
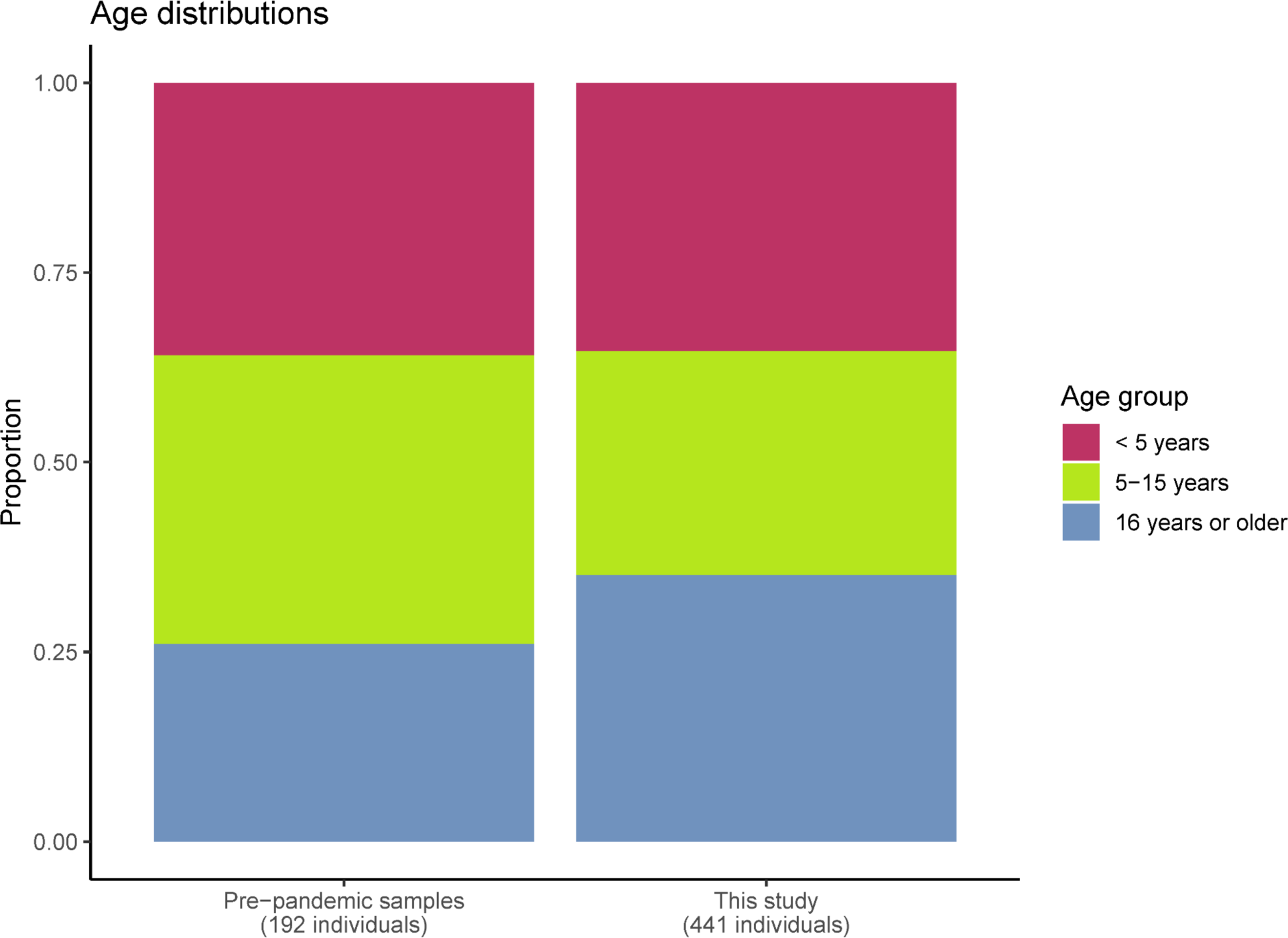
The age distribution of this study population, compared to the age distribution of the negative control samples used.

**Supplementary Figure 5:**
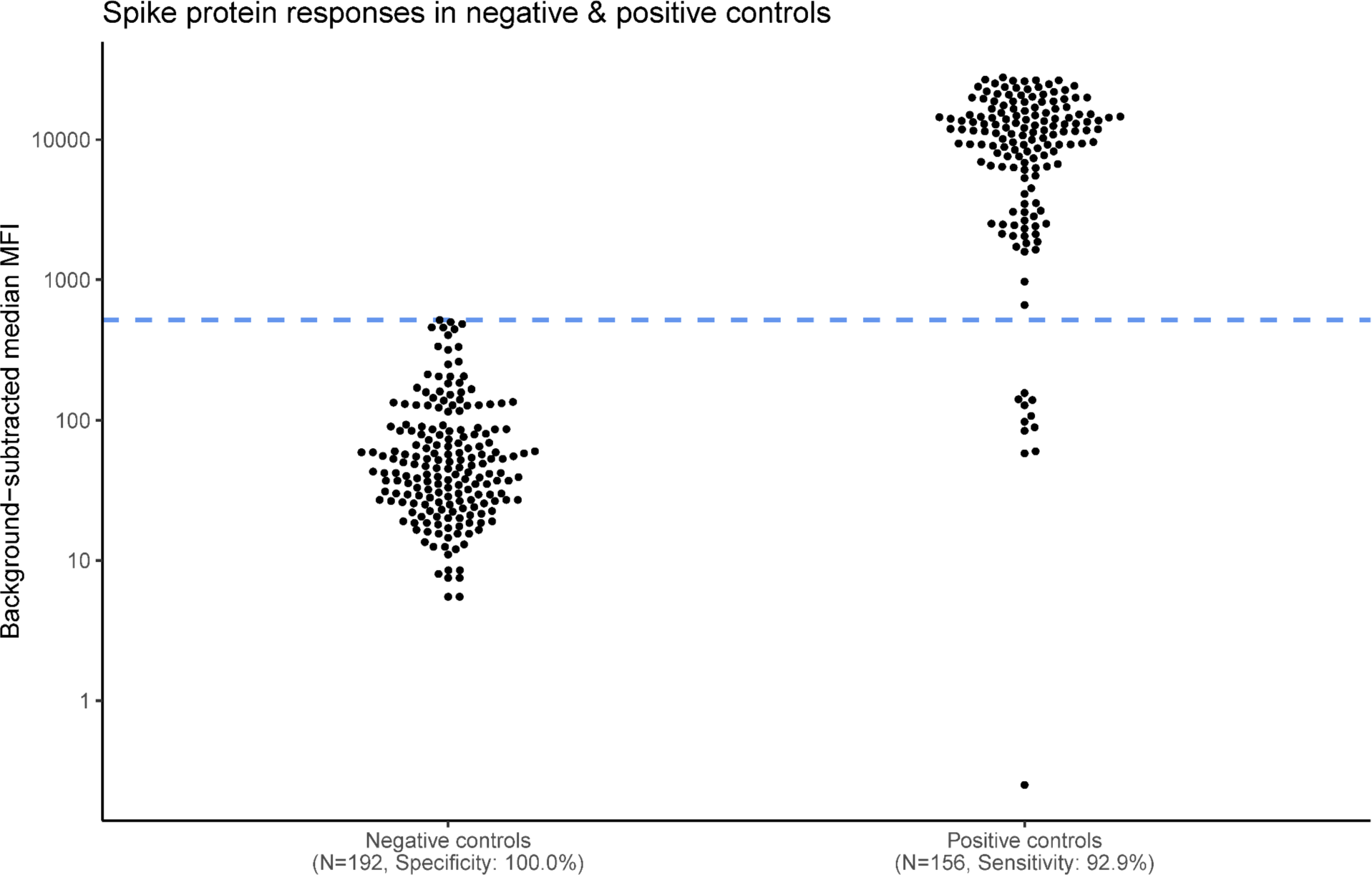
Spike protein antibody responses for SARS-CoV-2 negative control and positive control samples. The cutoff for seropositivity is shown in the blue dashed line (background-subtracted median MFI = 516).

**Supplementary Figure 6:**
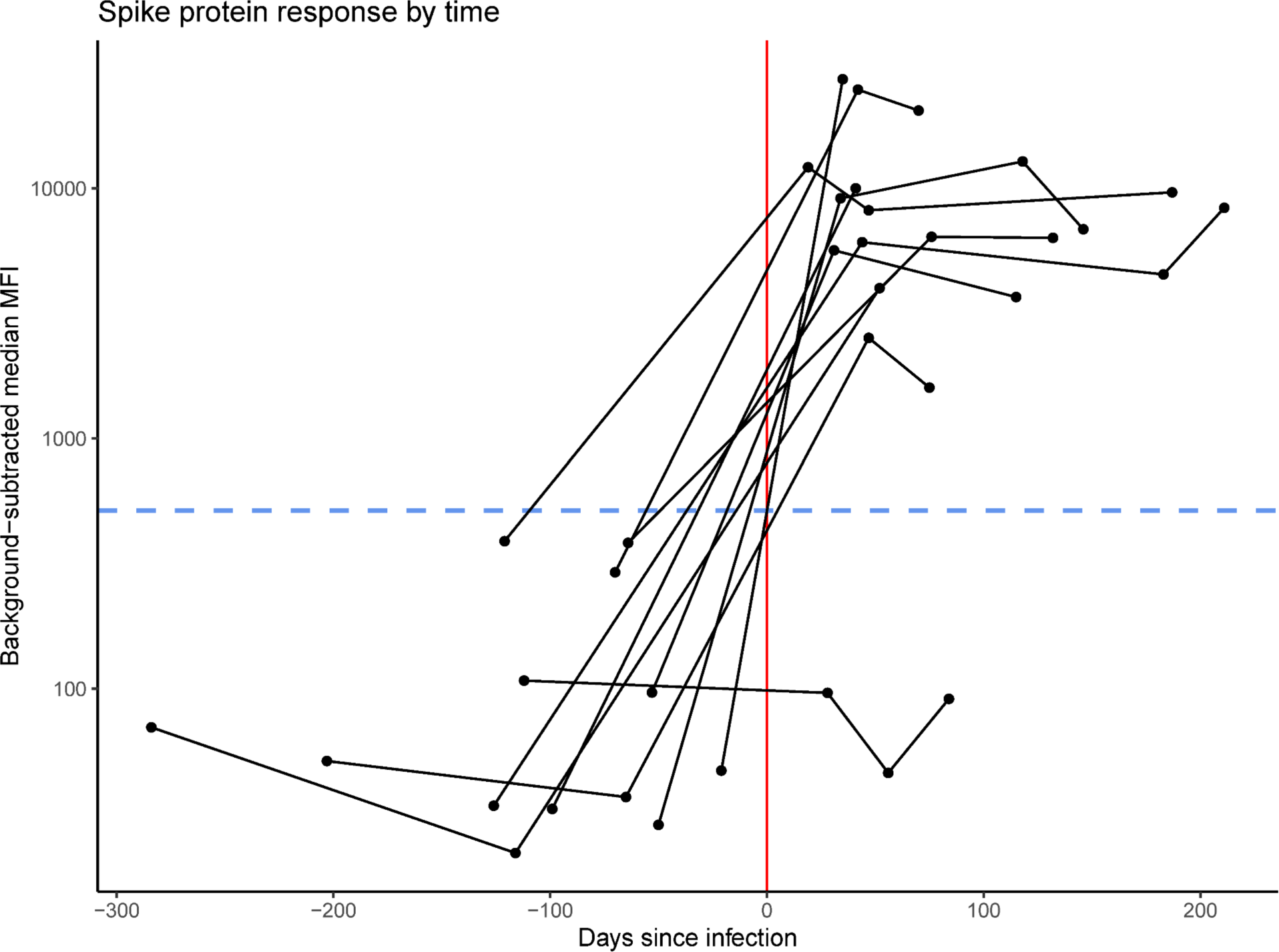
Longitudinal antibody kinetics for 11 participants in the cohort study who had confirmed SARS-CoV-2 infections. The x-axis represents days since infection and the y-axis represents the spike protein antibody response. The 11 infections occurred between February and July 2021. Samples from Round 4 of the serosurvey are omitted from this visualization to preclude boosting effects. The cutoff for seropositivity is shown in the blue dashed line (background-subtracted median MFI = 516).

**Supplementary Figure 7:**
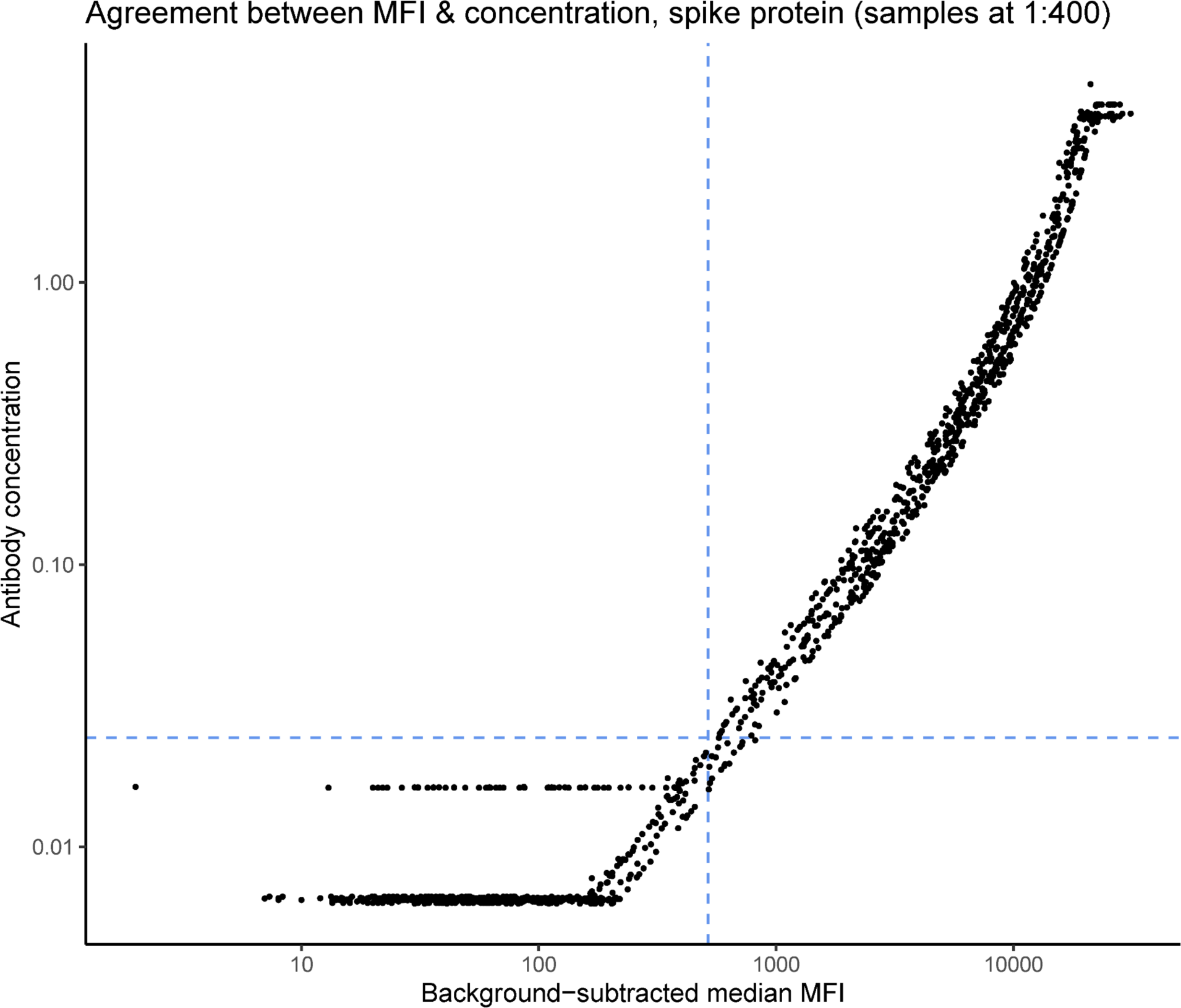
Scatterplot of agreement between measured MFIs and inferred antibody concentrations. Each point represents one of the 1,483 serosurvey samples tested at the primary concentration (1:400). Only 18 of 1,483 (1.2%) binary seropositivity results were not in agreement between MFIs and antibody concentrations, all of which were called positive using MFIs and negative using concentrations. The cutoff for seropositivity for each metric, determined based on the highest negative control, is shown in blue.

**Supplementary Figure 8:**
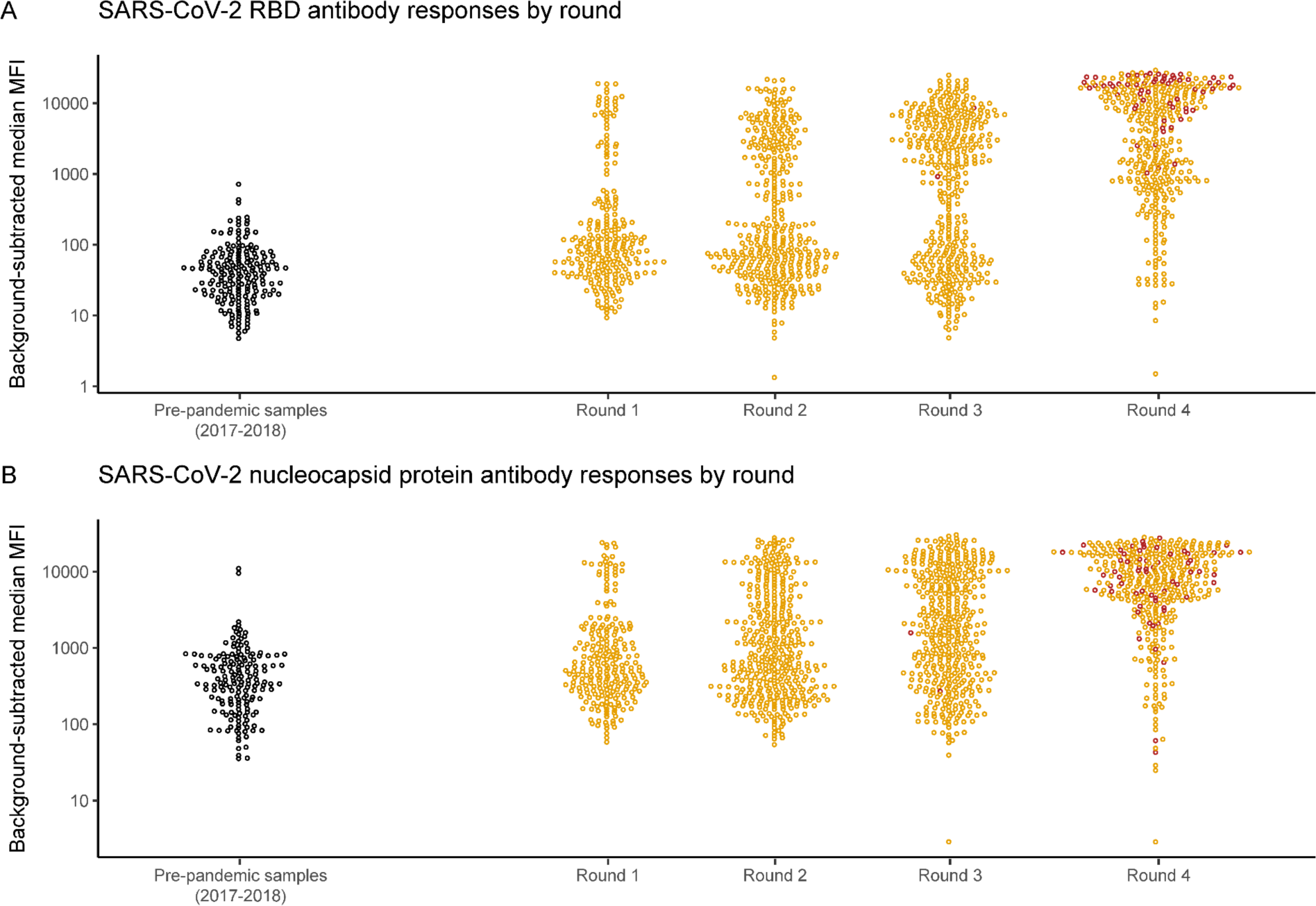
Antibody responses to additional SARS-CoV-2 antigens tested. Analogous to Figure 1B, here showing antibody responses to the **(A)** receptor-binding domain of the spike protein (RBD) and **(B)** nucleocapsid protein.

**Supplementary Figure 9:**
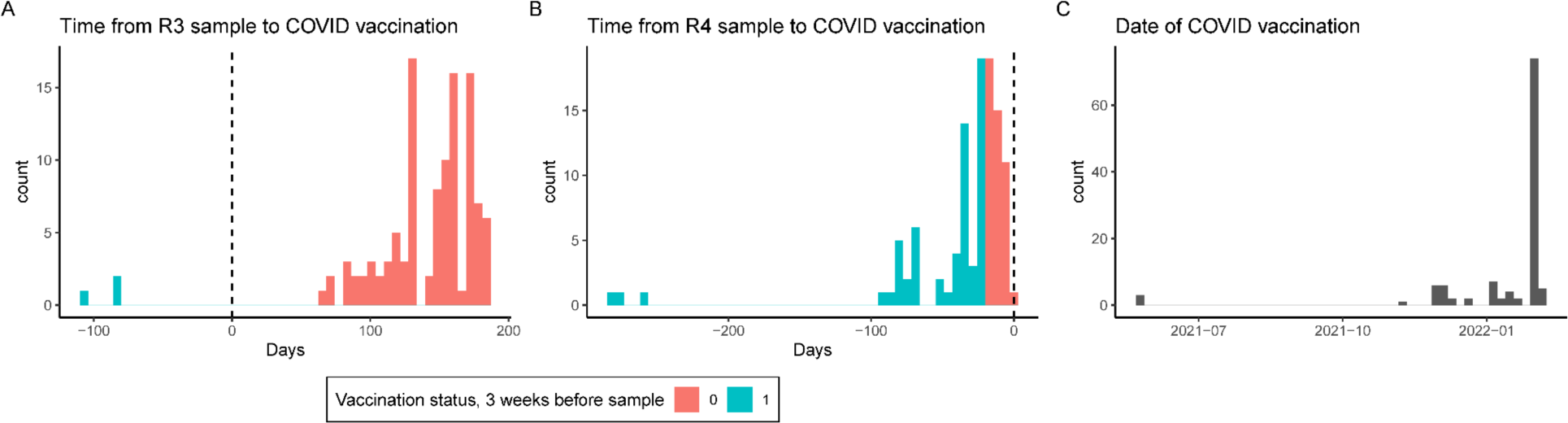
Timing of SARS-CoV-2 vaccination. Days between **(A)** Round 3 and **(B)** Round 4 of the serosurvey and vaccination date. The times are stratified by whether vaccination was received within the 3 weeks prior to sample collection (pink), or 3 or more weeks prior to sample collection (turquoise). Negative values of the x-axis indicate vaccination that occurred before sample collection (i.e., panel A indicates that all but 3 participants who received vaccination were vaccinated after their Round 3 serosurvey sample). **(C)** Dates of SARS-CoV-2 vaccination among study participants. Vaccines received included 83 Janssen/Johnson & Johnson, 12 AstraZeneca, 9 Moderna, and 1 Sinovac.

**Supplementary Figure 10:**
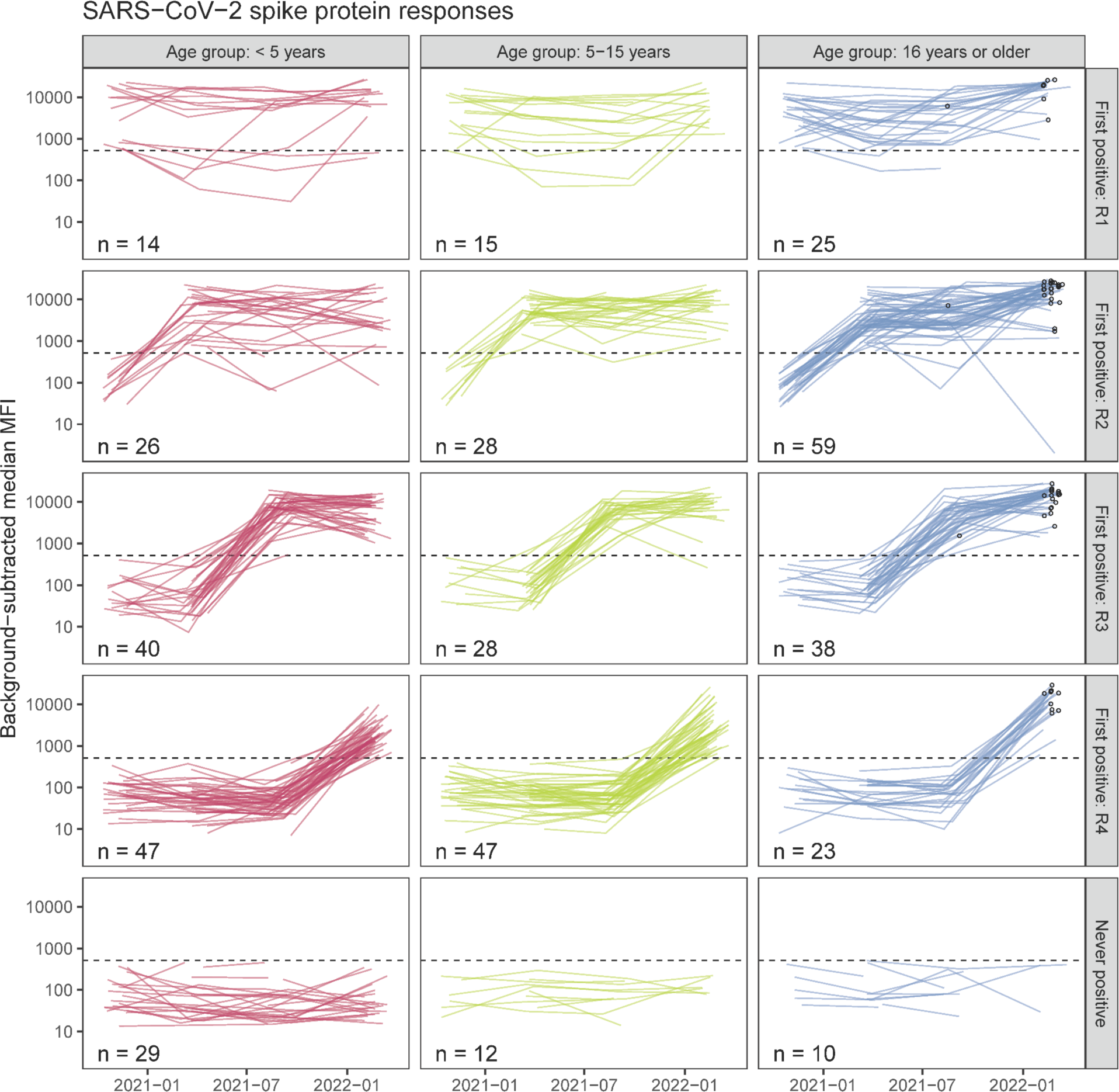
SARS-CoV-2 antibody kinetics, stratified by age group and by serosurvey round at which an individual’s antibody response was first positive. The kinetics for a single individual are shown by a line. The x-axis shows calendar time and the y-axis shows the background-subtracted median MFI of the spike protein antibody response. The bottom row indicates individuals whose antibody response was negative across the time period of this study. The value in the lower left of each panel indicates the number of individuals represented in that panel. The black circles indicate participants who had received SARS-coV-2 vaccination by the 3 weeks prior to the serosurvey sample collection.

**Supplementary Figure 11:**
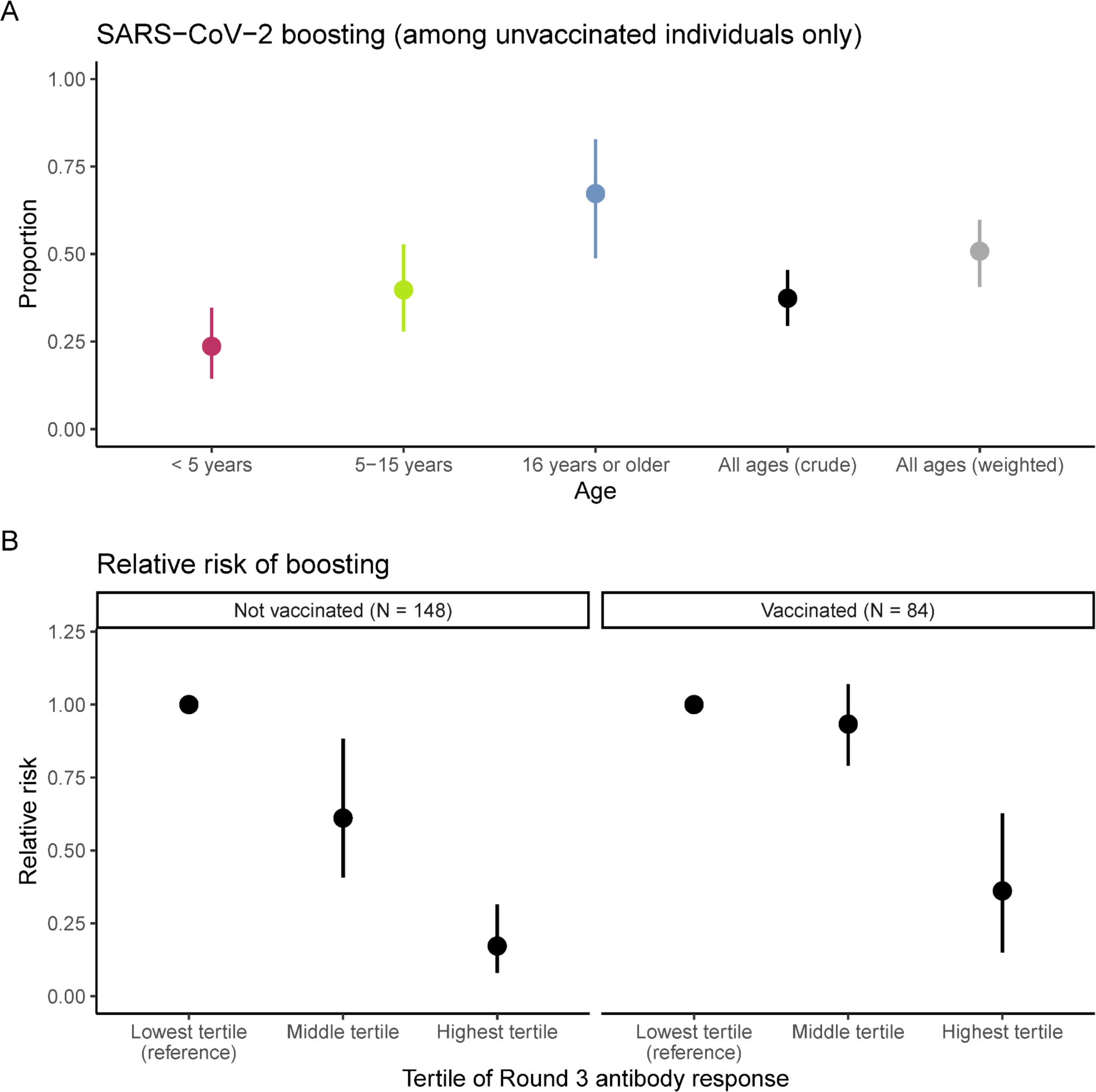
Antibody boosting by strata. **(A)** Probability of boosting among unvaccinated individuals only. The colors represent age group-specific estimates. The black values represent the crude estimates in the cohort. The gray values represent estimates weighted by the local age distribution using 2014 census data from the three parishes in Uganda in which study participants reside. **(B)** Relative risk of boosting by vaccination status at Round 4 and by baseline (Round 3) antibody tertile.

**Supplementary Figure 12:**
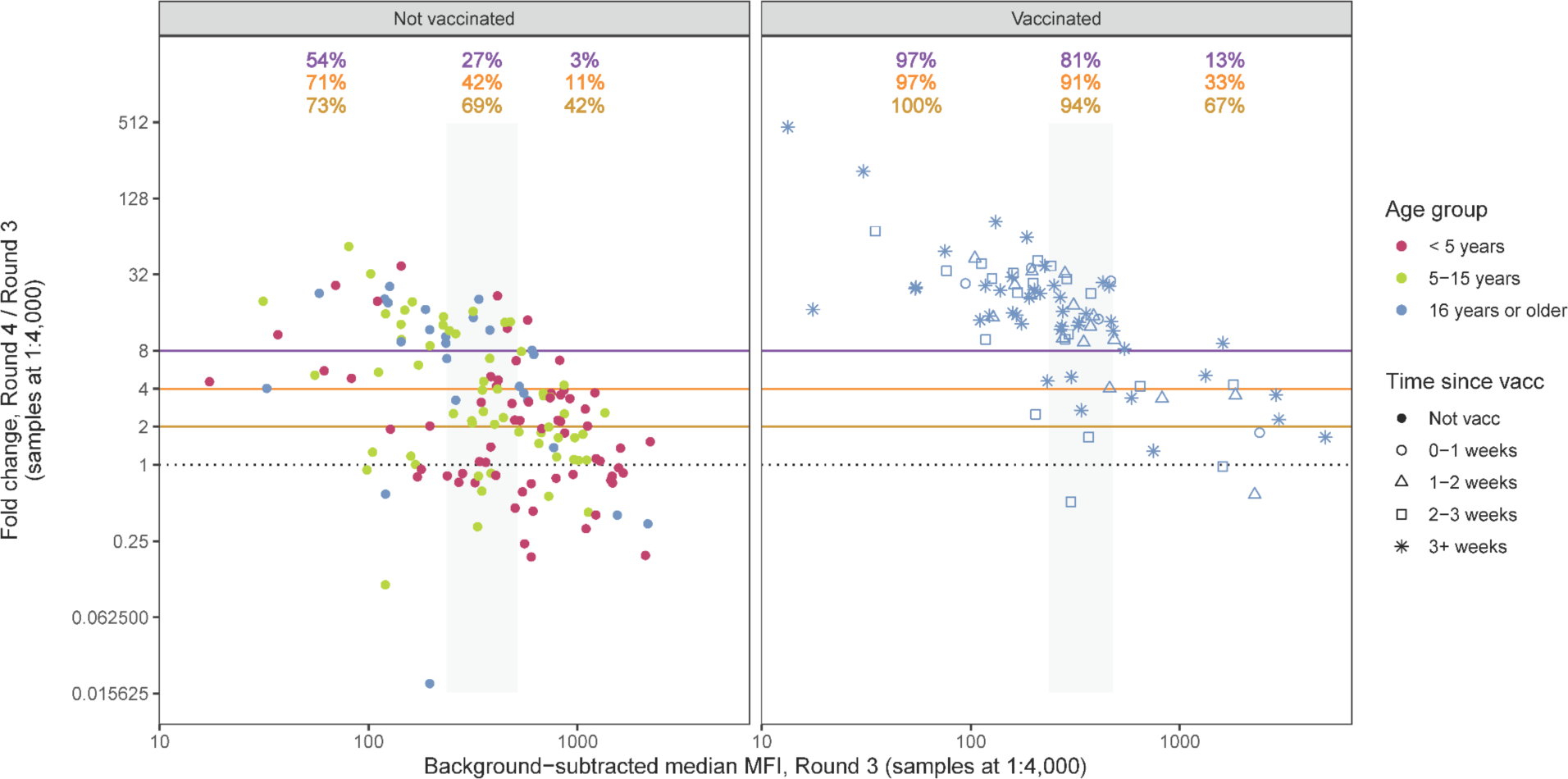
Sensitivity analyses for SARS-CoV-2 antibody boosting between Rounds 3 and 4 using alternative cutoffs. Analogous to Figure 3, here showing different cutoffs of boosting: ≥ 2 fold increase in MFI (gold), ≥ 4 fold increase in MFI (orange), and > 8 fold increase in MFI (purple) for spike protein. The Round 3 antibody response is shown on the x-axis, and the fold change between the Round 4 and Round 3 antibody response is shown on the y-axis. Participants were separated by vaccination status at Round 4 (panels) and by tertiles of Round 3 response (the second tertile is shown in the gray shaded rectangle). The proportion of individuals within each tertile that demonstrated antibody boosting is shown in gold, orange, and purple text at the top consistent with the respective colors of the lines representing cutoffs. The colors of the points represent age groups, and the shapes of the points represent binned time since SARS-CoV-2 vaccination at the Round 4 sample.

**Supplementary Figure 13:**
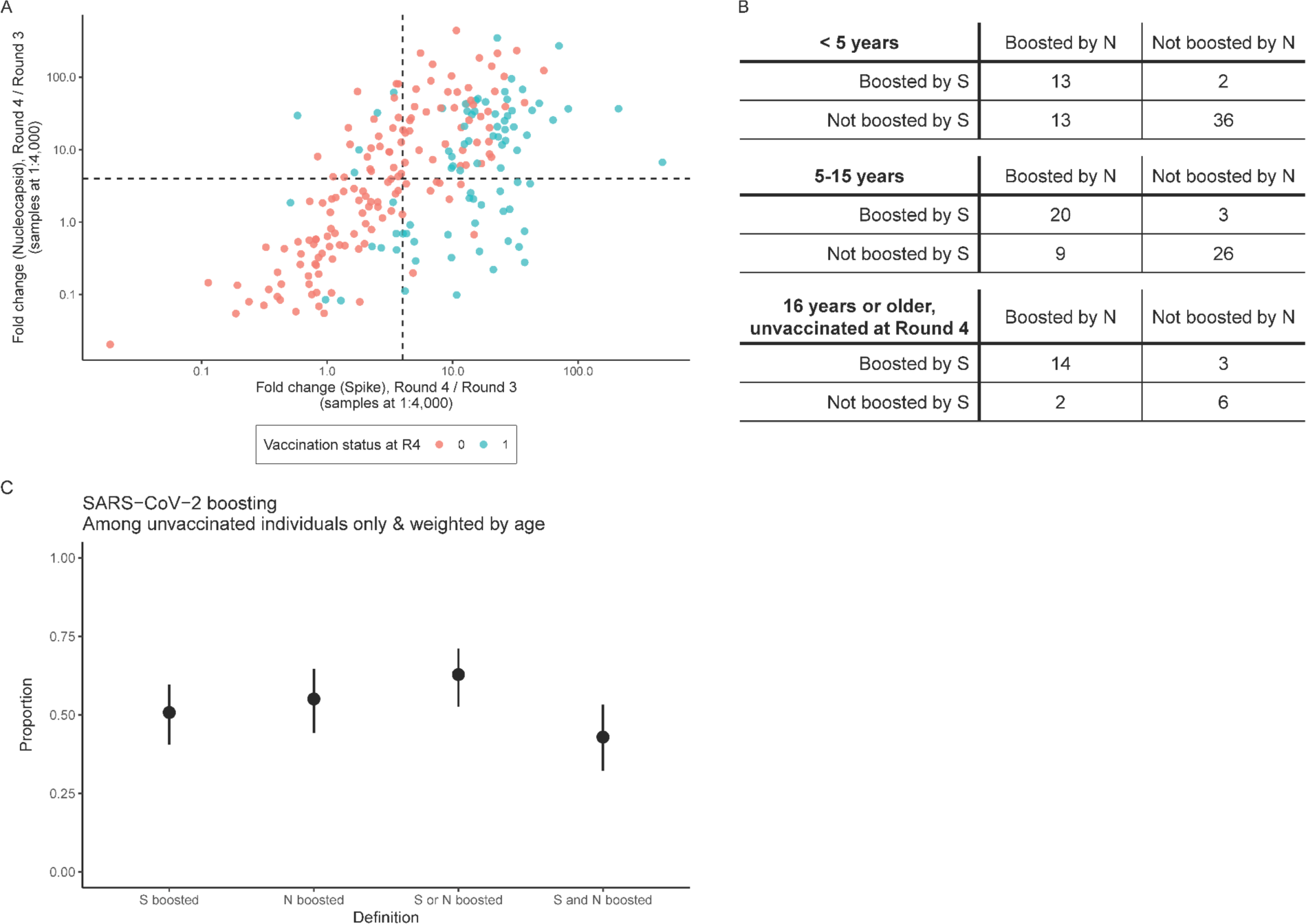
Sensitivity analyses for SARS-CoV-2 antibody boosting between Rounds 3 and 4 using nucleocapsid protein responses. **(A)** Fold change in spike (S, x-axis) or nucleocapsid (N, y-axis) between Round 3 and Round 4, for participants who were seropositive at Round 3 by spike. Note that vaccinated individuals on average had higher boosting to spike than nucleocapsid, consistent with inclusion of spike protein in the vaccine, but that some vaccinated individuals also had evidence of boosting with nucleocapsid, likely indicating concomitant re-infection. Four individuals with N responses that had less than the minimum of 50 beads per analyte were omitted. The dashed lines represent a 4 fold increase. **(B)** Frequency table of boosting of S and N responses by age group. Only participants who were seropositive at Round 3 by spike were considered at risk for boosting. Boosting was defined as a ≥ 4 fold increase in MFI for that protein between Round 3 and Round 4. (C) Age-weighted probability of boosting for each definition: boosting by S, boosting by N, boosting by S or N, boosting by S and N.

**Supplementary Figure 14:**
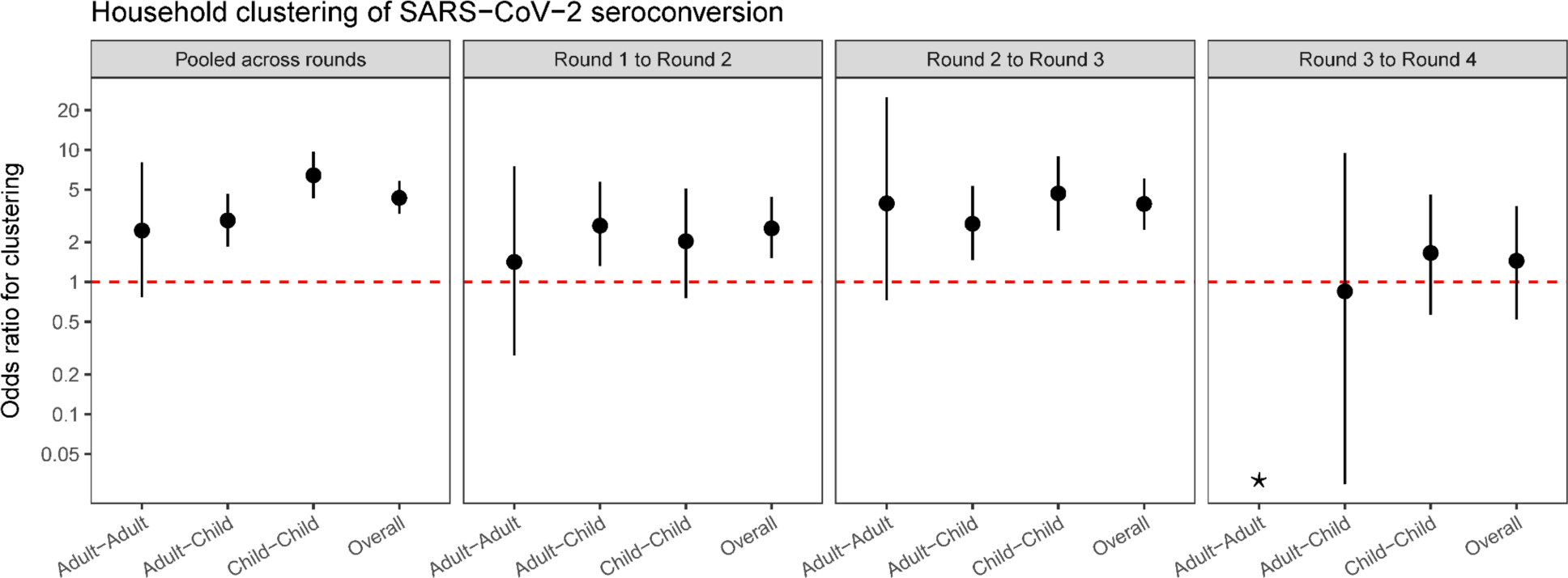
Pairwise odds ratios for clustering of SARS-CoV-2 seroconversions within households, by time period and by age category. The leftmost panel depicts results of pooling seroconversions across all intervals; the subsequent panels depict results stratified by interval. Individuals who had received SARS-CoV-2 vaccination by a serosurvey round are removed from the risk set for seroconversion. For each time period, we estimated within-age category and between-age category pairwise odds ratios for clustering within households (“Adult-Adult”, “Adult-Child”, “Child-Child”), as well as a single overall pairwise odds ratio for clustering within households (“Overall”). For this analysis, we defined children as individuals 15 years of age or younger, and adults are individuals 16 years of age or older. *A pairwise odds ratio for clustering within adults in households was not estimated for the interval between Round 3 and Round 4, as there were no households that had greater than 1 adult who was unvaccinated and susceptible. The dashed red line is for odds ratio = 1.

## Notes

### Competing Interest Statement

The authors have declared no competing interest.

