## Supplemental Information for "Reconstructing the SARS-CoV-2 epidemic in eastern Uganda through longitudinal serosurveillance in a malaria cohort"

### SUPPLEMENTARY TABLES

**Supplementary Table 1: Number of samples seropositive for SARS-CoV-2 by spike protein MFI and total number of samples tested, by serosurvey round and age group.**

| Serosurvey round | Age group | Seropositive | Total |
|---|---|---|---|
| 1 | < 5 years | 14 | 89 |
| 1 | 5-15 years | 15 | 69 |
| 1 | 16 years or older | 25 | 87 |
| 1 | All ages | 54 | 245 |
| 2 | < 5 years | 35 | 134 |
| 2 | 5-15 years | 40 | 126 |
| 2 | 16 years or older | 82 | 154 |
| 2 | All ages | 157 | 414 |
| 3 | < 5 years | 74 | 153 |
| 3 | 5-15 years | 68 | 129 |
| 3 | 16 years or older | 118 | 152 |
| 3 | 16 years or older, unvaccinated at Round 3 | 115 | 149 |
| 3 | All ages | 260 | 434 |
| 4 | < 5 years | 115 | 139 |
| 4 | 5-15 years, total | 107 | 114 |
| 4 | 5-15 years, unvaccinated at Round 4 | 107 | 113 |
| 4 | 16 years or older, total | 133 | 137 |
| 4 | 16 years or older, unvaccinated at Round 4 | 30 | 32 |
| 4 | All ages | 355 | 390 |

**Supplementary Table 2: Number of participants with SARS-CoV-2 seroconversion by spike MFI and total number of participants at risk for seroconversion, by interval and age group.** Individuals are removed from the risk set if they had received SARS-CoV-2 vaccination before or during the interval.

| Interval | Age group | Seroconverted during interval | Total at risk for seroconversion during interval |
|---|---|---|---|
| Round 1 to Round 2 | < 5 years | 13 | 68 |
| Round 1 to Round 2 | 5-15 years | 11 | 52 |
| Round 1 to Round 2 | 16 years or older | 30 | 62 |
| Round 1 to Round 2 | All ages | 54 | 182 |
| Round 2 to Round 3 | < 5 years | 32 | 97 |
| Round 2 to Round 3 | 5-15 years | 28 | 86 |
| Round 2 to Round 3 | 16 years or older | 37 | 69 |
| Round 2 to Round 3 | All ages | 97 | 252 |
| Round 3 to Round 4 | < 5 years | 49 | 72 |
| Round 3 to Round 4 | 5-15 years | 50 | 55 |
| Round 3 to Round 4 | 16 years or older | 6 | 7 |
| Round 3 to Round 4 | All ages | 105 | 134 |

**Supplementary Table 3: Number of participants with SARS-CoV-2 boosting by spike MFI and total number of participants at risk for boosting, by interval and age group.** Only participants who were seropositive at Round 3 were considered at risk for boosting. Boosting was defined as a ≥ 4 fold increase in MFI between Round 3 and Round 4.

| Age group | Vaccination status | Boosted during interval | Total at risk for boosting during interval |
|---|---|---|---|
| < 5 years | Unvaccinated at Round 4 | 15 | 65 |
| 5-15 years | Unvaccinated at Round 4 | 23 | 58 |
| 16 years or older | Unvaccinated at Round 4 | 17 | 25 |
| 16 years or older | Vaccinated between Round 3 and Round 4 | 68 | 81 |
| 16 years or older | Vaccinated between Round 2 and Round 3 | 2 | 3 |
| All ages | -- | 125 | 232 |

**Supplementary Table 4: Symptoms and diagnoses associated with SARS-CoV-2 seroconversion.** Diarrhea, fever, muscle aches, and other diagnostic categories assessed were not associated with seroconversion.

| Characteristic | OR[1] | 95% CI[1] | p-value |
|---|---|---|---|
| **Cough** | 1.44 | 1.02, 2.02 | **0.037** |
| **Headache** | 2.04 | 1.43, 2.92 | **<0.001** |
| **Fatigue** | 1.82 | 1.04, 3.21 | **0.037** |
| **Total number sick visits** | 1.17 | 1.07, 1.29 | **<0.001** |
| **Any URTI diagnosis** | 1.86 | 1.21, 2.87 | **0.005** |
| **Total number URTI diagnoses** | 1.67 | 1.18, 2.35 | **0.003** |

[1]OR = Odds Ratio, CI = Confidence Interval

**Supplementary Table 5: Risk factors for antibody boosting & associations with malaria.**

| Characteristic | OR[1] | 95% CI[1] | p-value |
|---|---|---|---|
| **Age Category** | | | |
| < 5 years | — | — | |
| 5-15 years | 2.19 | 1.01, 4.86 | **0.049** |
| **Gender** | | | |
| Female | — | — | |
| Male | 0.93 | 0.43, 2.01 | 0.86 |
| **Wealth Tertile** | | | |
| Lowest | — | — | |
| Middle | 0.76 | 0.28, 2.17 | 0.61 |
| Highest | 0.42 | 0.14, 1.24 | 0.11 |
| **Housing Type** | | | |
| Modern | — | — | |
| Traditional | 0.46 | 0.21, 1.01 | 0.053 |
| **Sanitation** | | | |
| Uncovered pit latrine or no facility | — | — | |
| VIP or covered pit latrine | 1.17 | 0.50, 2.67 | 0.71 |
| **Number of people in household** | 1.26 | 0.92, 1.77 | 0.16 |
| **Malaria incidence** | 0.95 | 0.79, 1.12 | 0.60 |
| **Any asymptomatic parasitemia** | 1.60 | 0.74, 3.49 | 0.23 |
| **Number of episodes of asymptomatic parasitemia** | 1.37 | 1.05, 1.81 | **0.023** |

[1]OR = Odds Ratio, CI = Confidence Interval

### SUPPLEMENTARY FIGURES

**Supplementary Figure 1: District-level weekly SARS-CoV-2 case counts in eastern Uganda, compared to in the country overall.** In blue and on the left y-axis: aggregated weekly SARS-CoV-2 positive results from the 20 districts in eastern Uganda highlighted on the inset map (Budaka, Bugiri, Bugweri, Buikwe, Bukwo, Bulambuli, Busia, Butaleja, Butebo, Iganga, Jinja, Kaliro, Kibuku, Luuka, Mayuge, Mbale, Namisindwa, Namutumba, Pallisa, and Sironko). On the right y-axis: reported daily new COVID-19 cases in all of Uganda, replicated from Figure 1A.

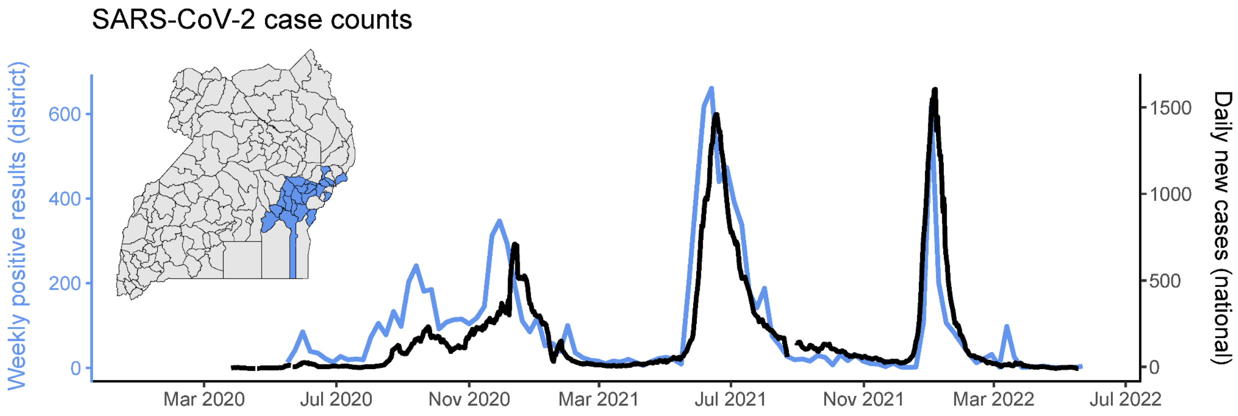

**Supplementary Figure 2: Standard dilution series used for assay normalization.** The x-axis represents the log antibody concentration and the y-axis represents the median MFI value of the spike protein response. The black points indicate the serial dilution (inverse of concentration) of the standards. Each line represents a plate, with 32 plates tested in total. The background response (average of blank wells) for each plate is shown in blue.

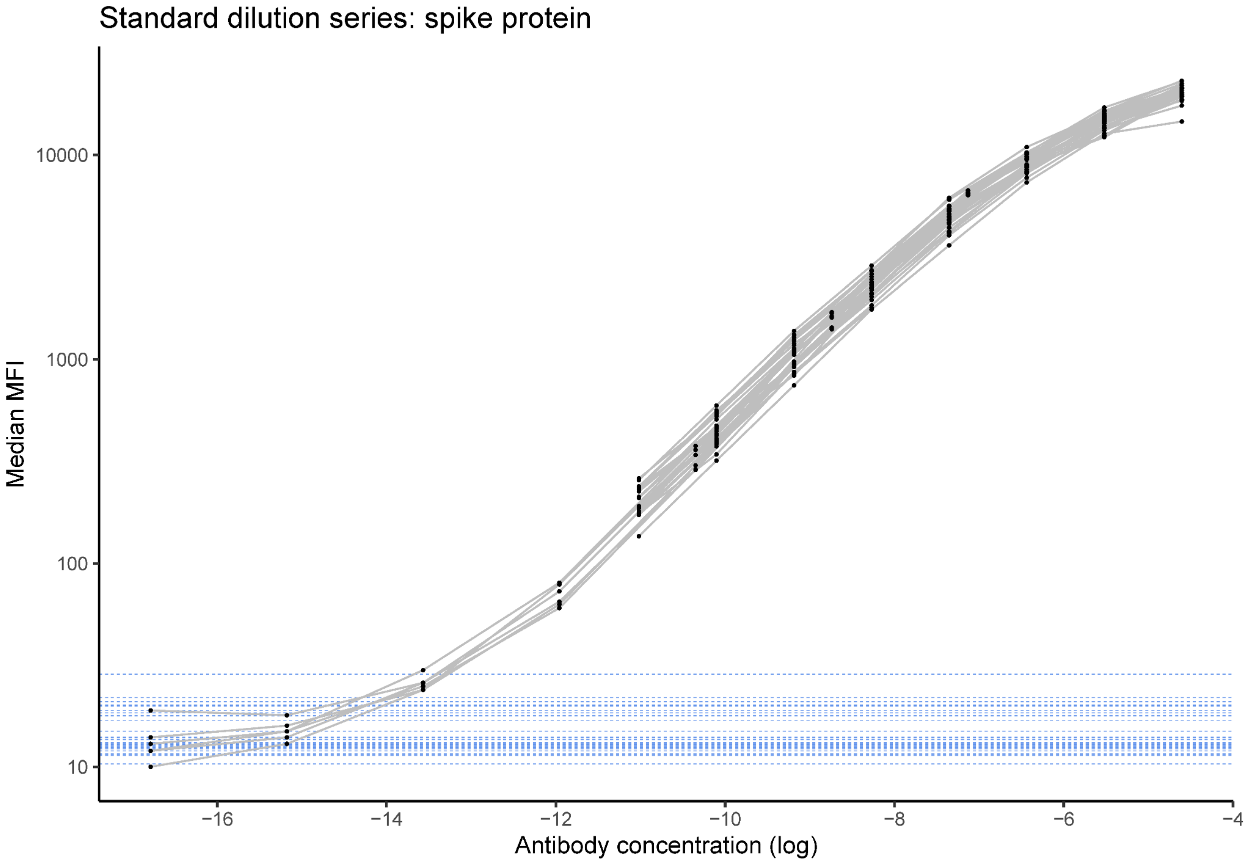

**Supplementary Figure 3: Replicability of the Luminex assay.** Scatterplots of spike protein **(A)** MFIs and **(B)** antibody concentrations for 50 samples tested in replicate. The cutoff for seropositivity for each metric, determined based on the highest negative control, is shown in blue. On Panel B, samples that were below the limit of quantitation for antibody concentrations (i.e., below the lowest standard dilution on the plate) are shown in red.

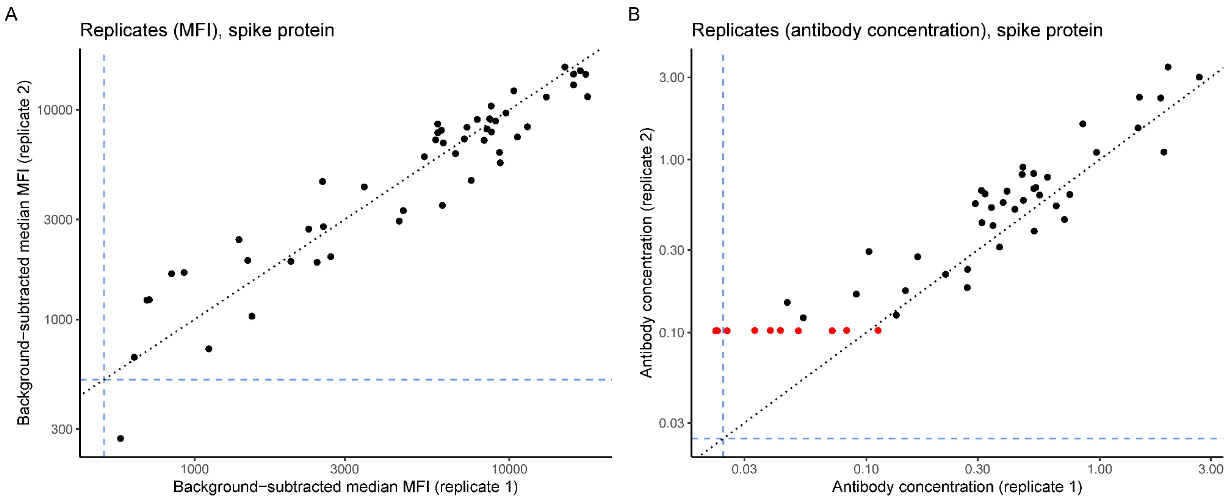

**Supplementary Figure 4: The age distribution of this study population, compared to the age distribution of the negative control samples used.**

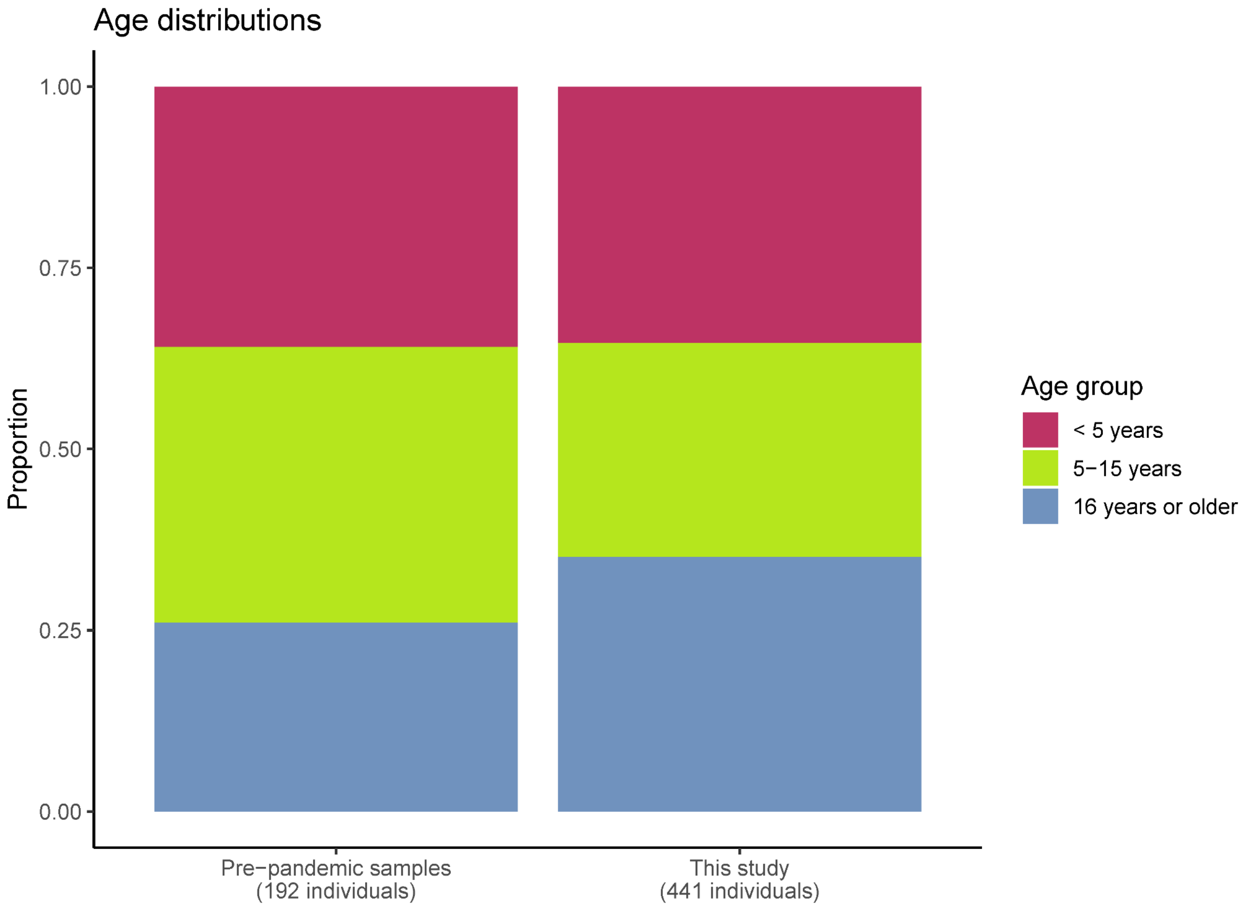

**Supplementary Figure 5: Spike protein antibody responses for SARS-CoV-2 negative control and positive control samples.** The cutoff for seropositivity is shown in the blue dashed line (background-subtracted median MFI = 516).

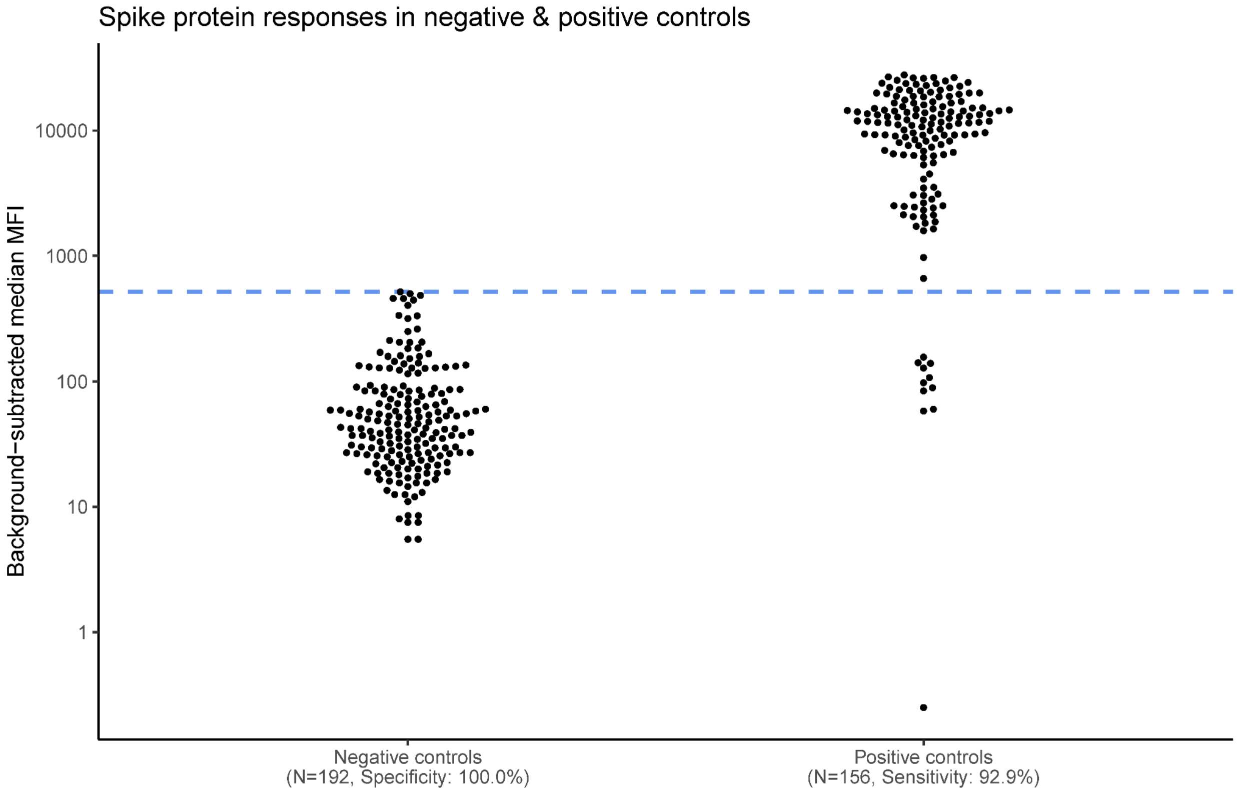

**Supplementary Figure 6: Longitudinal antibody kinetics for 11 participants in the cohort study who had confirmed SARS-CoV-2 infections.** The x-axis represents days since infection and the y-axis represents the spike protein antibody response. The 11 infections occurred between February and July 2021. Samples from Round 4 of the serosurvey are omitted from this visualization to preclude boosting effects. The cutoff for seropositivity is shown in the blue dashed line (background-subtracted median MFI = 516).

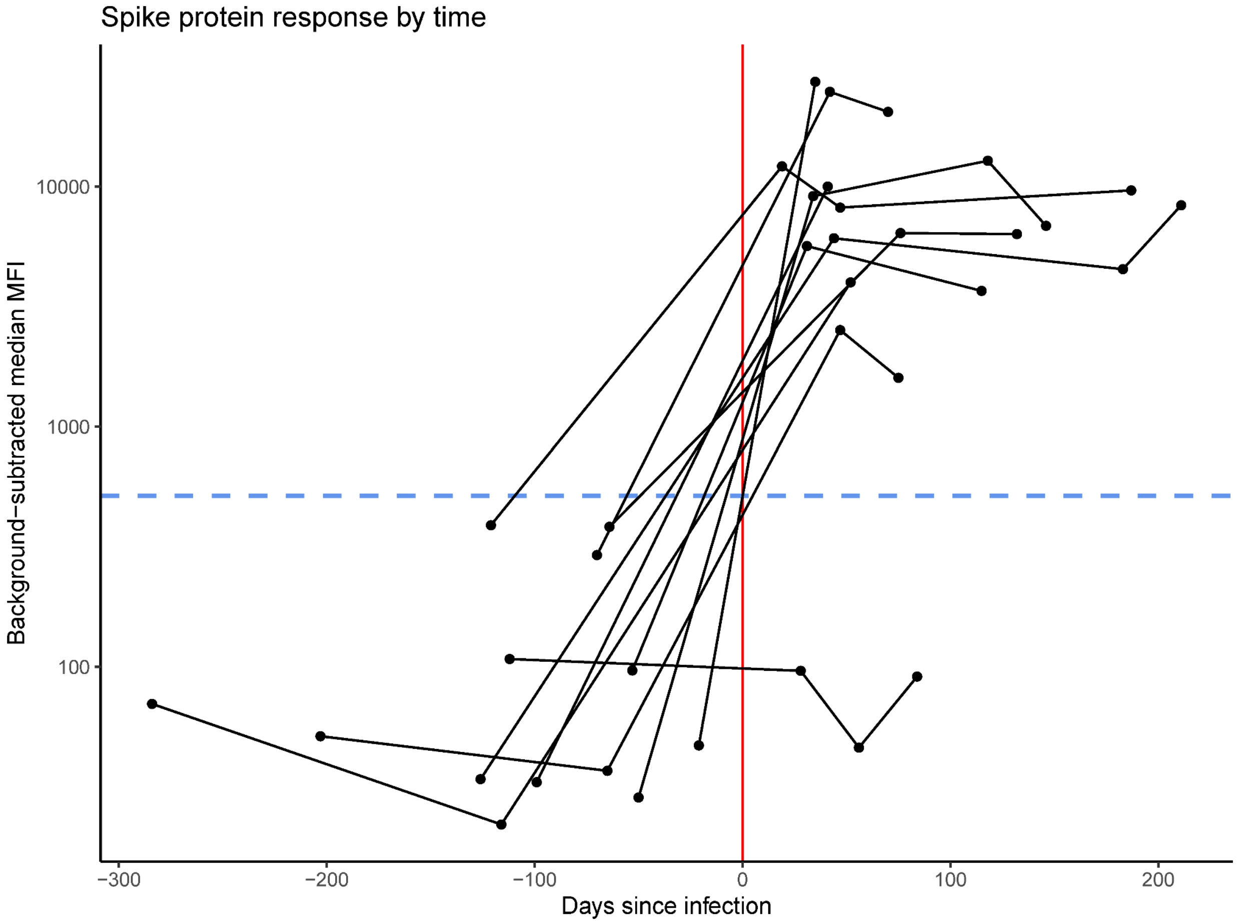

**Supplementary Figure 7: Scatterplot of agreement between measured MFIs and inferred antibody concentrations.** Each point represents one of the 1,483 serosurvey samples tested at the primary concentration (1:400). Only 18 of 1,483 (1.2%) binary seropositivity results were not in agreement between MFIs and antibody concentrations, all of which were called positive using MFIs and negative using concentrations. The cutoff for seropositivity for each metric, determined based on the highest negative control, is shown in blue.

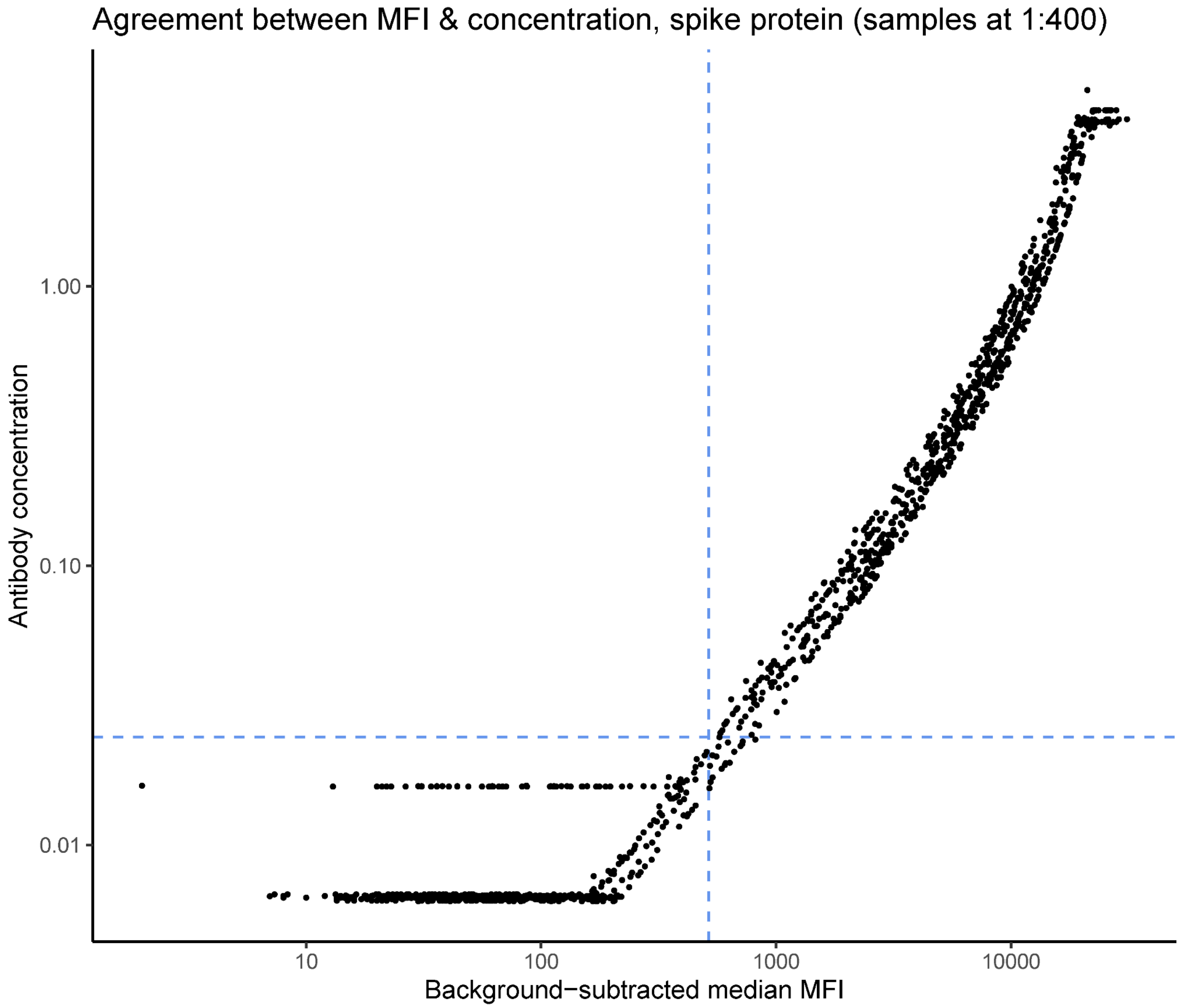

**Supplementary Figure 8: Antibody responses to additional SARS-CoV-2 antigens tested.** Analogous to Figure 1B, here showing antibody responses to the **(A)** receptor-binding domain of the spike protein (RBD) and **(B)** nucleocapsid protein.

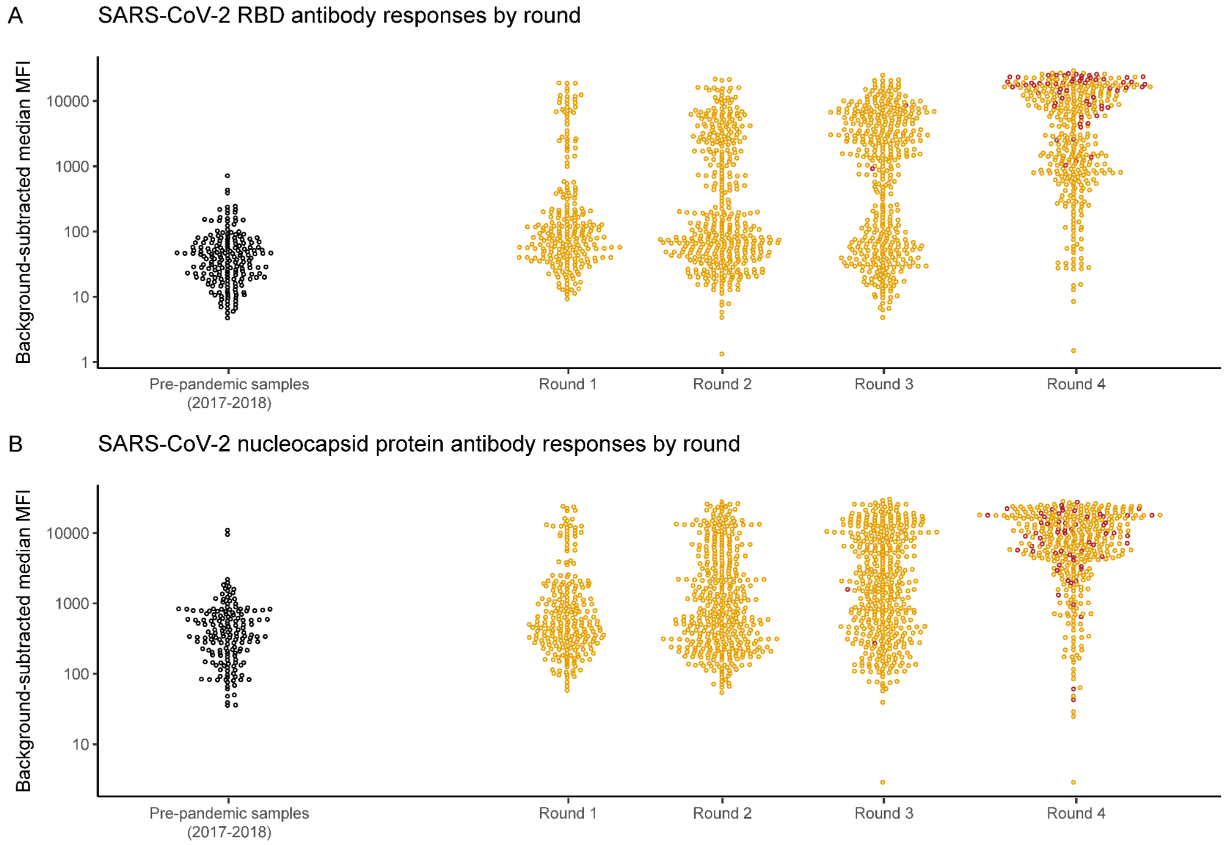

**Supplementary Figure 9: Timing of SARS-CoV-2 vaccination.** Days between **(A)** Round 3 and **(B)** Round 4 of the serosurvey and vaccination date. The times are stratified by whether vaccination was received within the 3 weeks prior to sample collection (pink), or 3 or more weeks prior to sample collection (turquoise). Negative values of the x-axis indicate vaccination that occurred before sample collection (i.e., panel A indicates that all but 3 participants who received vaccination were vaccinated after their Round 3 serosurvey sample). **(C)** Dates of SARS-CoV-2 vaccination among study participants. Vaccines received included 83 Janssen/Johnson & Johnson, 12 AstraZeneca, 9 Moderna, and 1 Sinovac.

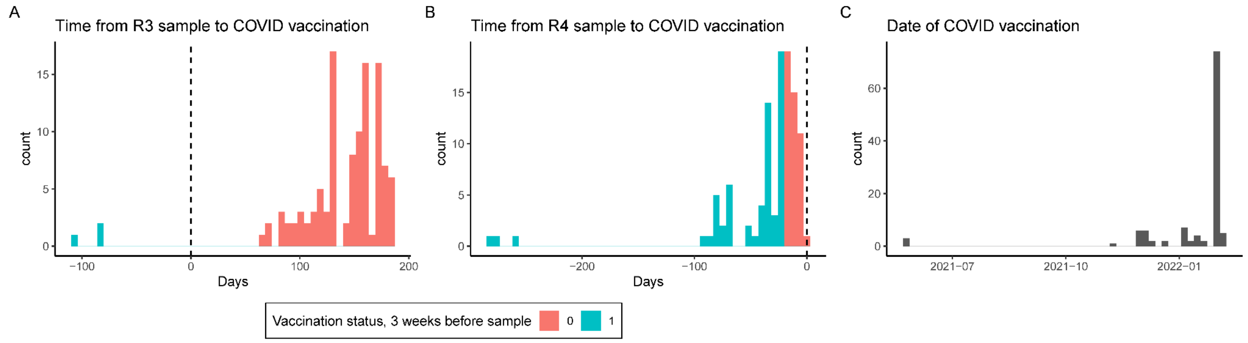

**Supplementary Figure 10: SARS-CoV-2 antibody kinetics, stratified by age group and by serosurvey round at which an individual's antibody response was first positive.** The kinetics for a single individual are shown by a line. The x-axis shows calendar time and the y-axis shows the background-subtracted median MFI of the spike protein antibody response. The bottom row indicates individuals whose antibody response was negative across the time period of this study. The value in the lower left of each panel indicates the number of individuals represented in that panel. The black circles indicate participants who had received SARS-coV-2 vaccination by the 3 weeks prior to the serosurvey sample collection.

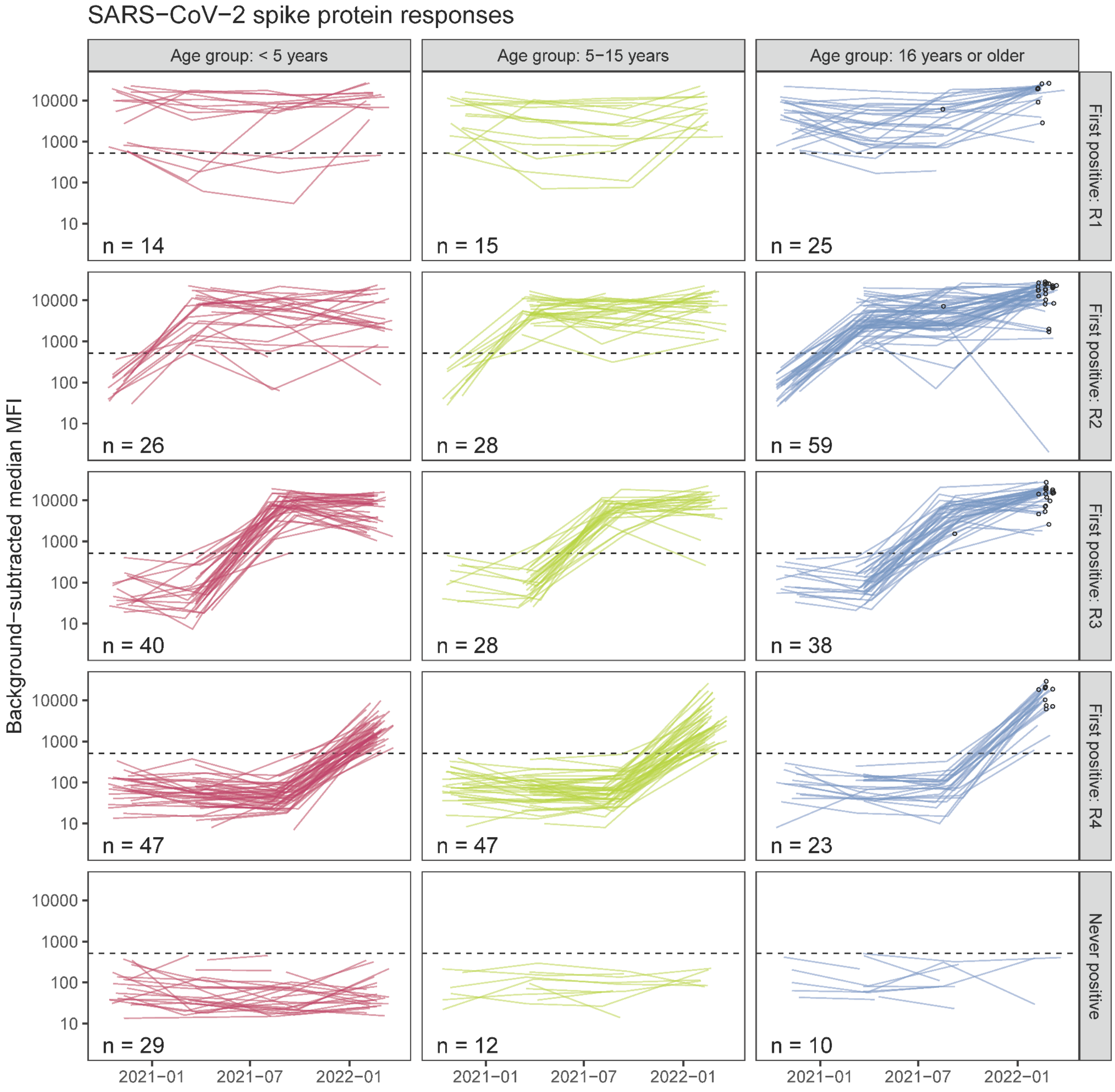

**Supplementary Figure 11: Antibody boosting by strata. (A)** Probability of boosting among unvaccinated individuals only. The colors represent age group-specific estimates. The black values represent the crude estimates in the cohort. The gray values represent estimates weighted by the local age distribution using 2014 census data from the three parishes in Uganda in which study participants reside. **(B)** Relative risk of boosting by vaccination status at Round 4 and by baseline (Round 3) antibody tertile.

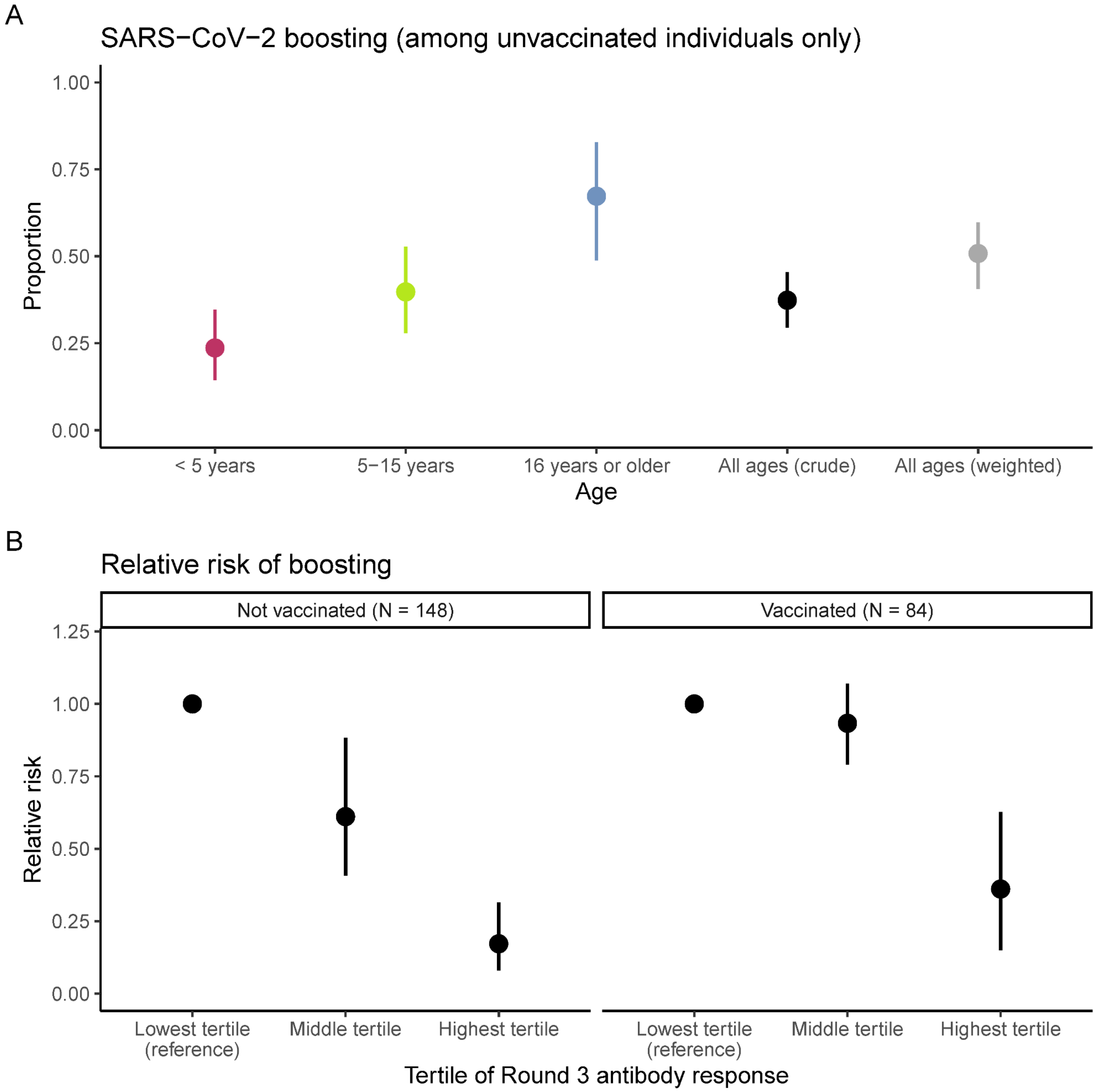

**Supplementary Figure 12: Sensitivity analyses for SARS-CoV-2 antibody boosting between Rounds 3 and 4 using alternative cutoffs.** Analogous to Figure 3, here showing different cutoffs of boosting: ≥ 2 fold increase in MFI (gold), ≥ 4 fold increase in MFI (orange), and ≥ 8 fold increase in MFI (purple) for spike protein. The Round 3 antibody response is shown on the x-axis, and the fold change between the Round 4 and Round 3 antibody response is shown on the y-axis. Participants were separated by vaccination status at Round 4 (panels) and by tertiles of Round 3 response (the second tertile is shown in the gray shaded rectangle). The proportion of individuals within each tertile that demonstrated antibody boosting is shown in gold, orange, and purple text at the top consistent with the respective colors of the lines representing cutoffs. The colors of the points represent age groups, and the shapes of the points represent binned time since SARS-CoV-2 vaccination at the Round 4 sample.

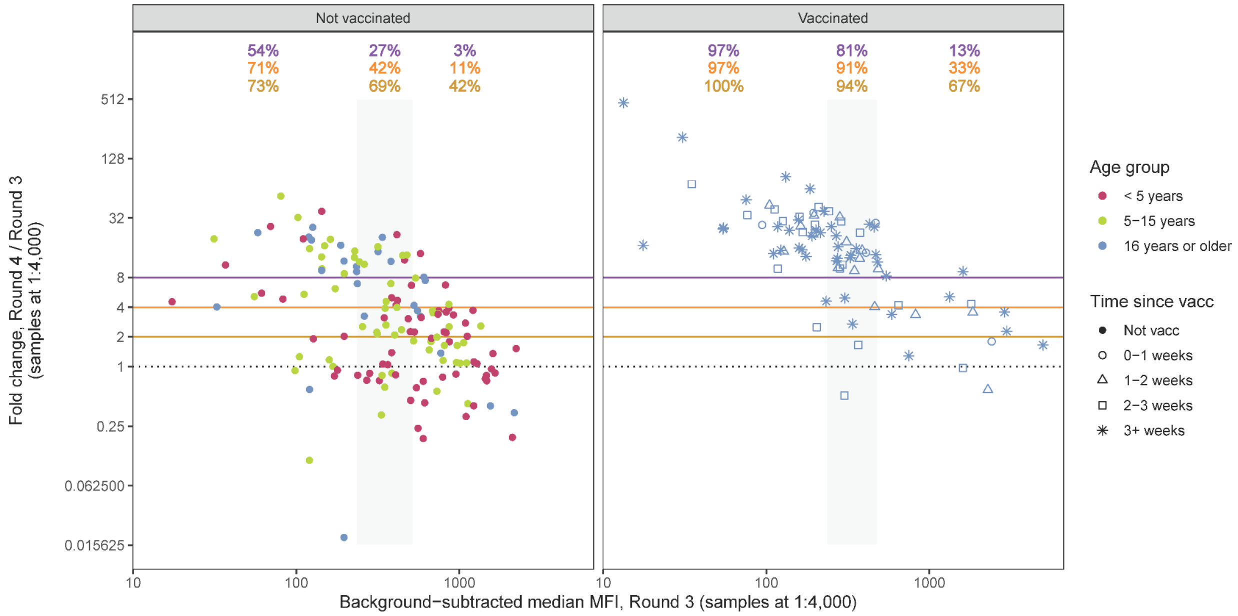

**Supplementary Figure 13: Sensitivity analyses for SARS-CoV-2 antibody boosting between Rounds 3 and 4 using nucleocapsid protein responses. (A)** Fold change in spike (S, x-axis) or nucleocapsid (N, y-axis) between Round 3 and Round 4, for participants who were seropositive at Round 3 by spike. Note that vaccinated individuals on average had higher boosting to spike than nucleocapsid, consistent with inclusion of spike protein in the vaccine, but that some vaccinated individuals also had evidence of boosting with nucleocapsid, likely indicating concomitant re-infection. Four individuals with N responses that had less than the minimum of 50 beads per analyte were omitted. The dashed lines represent a 4 fold increase. **(B)** Frequency table of boosting of S and N responses by age group. Only participants who were seropositive at Round 3 by spike were considered at risk for boosting. Boosting was defined as a ≥ 4 fold increase in MFI for that protein between Round 3 and Round 4. **(C)** Age-weighted probability of boosting for each definition: boosting by S, boosting by N, boosting by S or N, boosting by S and N.

A

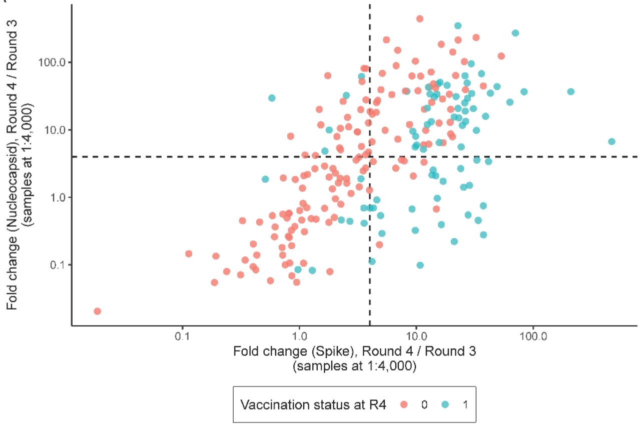


B

| **< 5 years** | Boosted by N | Not boosted by N |
|---|---|---|
| Boosted by S | 13 | 2 |
| Not boosted by S | 13 | 36 |

| **5-15 years** | Boosted by N | Not boosted by N |
|---|---|---|
| Boosted by S | 20 | 3 |
| Not boosted by S | 9 | 26 |

| **16 years or older, unvaccinated at Round 4** | Boosted by N | Not boosted by N |
|---|---|---|
| Boosted by S | 14 | 3 |
| Not boosted by S | 2 | 6 |

C

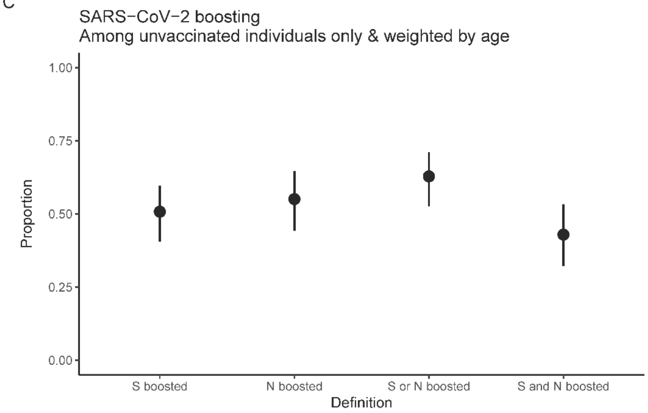

**Supplementary Figure 14: Pairwise odds ratios for clustering of SARS-CoV-2 seroconversions within households, by time period and by age category.** The leftmost panel depicts results of pooling seroconversions across all intervals; the subsequent panels depict results stratified by interval. Individuals who had received SARS-CoV-2 vaccination by a serosurvey round are removed from the risk set for seroconversion. For each time period, we estimated within-age category and between-age category pairwise odds ratios for clustering within households (“Adult-Adult”, “Adult-Child”, “Child-Child”), as well as a single overall pairwise odds ratio for clustering within households (“Overall”). For this analysis, we defined children as individuals 15 years of age or younger, and adults are individuals 16 years of age or older. *A pairwise odds ratio for clustering within adults in households was not estimated for the interval between Round 3 and Round 4, as there were no households that had greater than 1 adult who was unvaccinated and susceptible. The dashed red line is for odds ratio = 1.

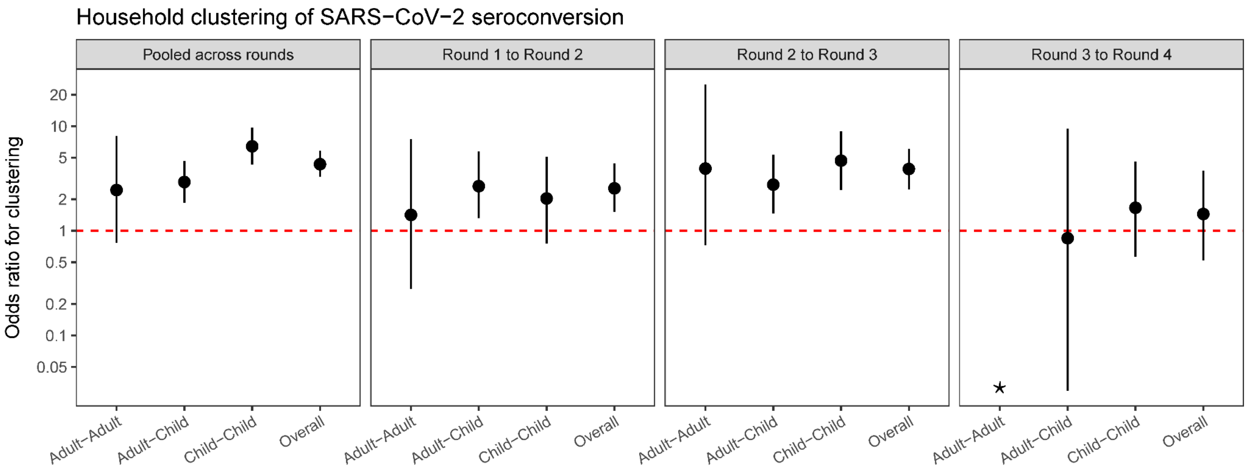
